# Artificial intelligence automation of echocardiographic measurements

**DOI:** 10.1101/2025.03.18.25324215

**Authors:** Yuki Sahashi, Hirotaka Ieki, Victoria Yuan, Matthew Christensen, Milos Vukadinovic, Christina Binder-Rodriguez, Justin Rhee, James Y. Zou, Bryan He, Paul Cheng, David Ouyang

## Abstract

**Background:** Accurate measurement of echocardiographic parameters is crucial for the diagnosis of cardiovascular disease and tracking of change over time, however manual assessment is time-consuming and can be imprecise. Artificial intelligence (AI) has the potential to reduce clinician burden by automating the time-intensive task of comprehensive measurement of echocardiographic parameters.

**Methods:** We developed and validated open-sourced deep learning semantic segmentation models for the automated measurement of 18 anatomic and Doppler measurements in echocardiography. The outputs of segmentation models were compared to sonographer measurements from two institutions to access accuracy and precision.

**Results:** We utilized 877,983 echocardiographic measurements from 155,215 studies from Cedars-Sinai Medical Center (CSMC) to develop EchoNet-Measurements, an open-source deep learning model for echocardiographic annotation. The models demonstrated a good correlation when compared with sonographer measurements from held-out data from CSMC and an independent external validation dataset from Stanford Healthcare (SHC). Measurements across all nine B-mode and nine Doppler measurements had high accuracy (an overall R^2^ of 0.967 (0.965 – 0.970) in the held-out CSMC dataset and 0.987 (0.984 – 0.989) in the SHC dataset). When evaluated end-to-end on a temporally distinct 2,103 studies at CSMC, EchoNet-Measurements performed well an overall R2 of 0.981 (0.976 – 0.984). Performance was consistent across patient characteristics including sex and atrial fibrillation status.

**Conclusion:** EchoNet-Measurement achieves high accuracy in automated echocardiographic measurement that is comparable to expert sonographers. This open-source model provides the foundation for future developments in AI applied to echocardiography.

**Clinical Perspective:** *What Is New?:*

- We developed EchoNet-Measurements, the first publicly available deep learning framework for comprehensive automated echocardiographic measurements.
- We assessed the performance of EchoNet-Measurements, showing good precision and accuracy compared to human sonographers and cardiologists across multiple healthcare systems.

*What Are the Clinical Implications?:*

- Deep-learning automated echocardiographic measurements can be conducted in a fraction of a second, reducing the time burden on sonographers and standardizing measurements, and potentially enhance reproducibility and diagnostic reliability.
- This open-source model provides broad opportunities for widespread adoption in both clinical use and research, including in resource-limited settings.

## Introduction

Accurate assessment and diagnosis of cardiovascular disease requires detailed and reproducible assessment of cardiac dimensions and function. Echocardiography, the most common and readily available modality of cardiac imaging, enables frequent and rapid evaluation and surveillance of disease. However, echocardiographic measurements require detailed manual measurements and even expert evaluation has considerable variability^1,2^. Given the large number of relevant and important echocardiographic parameters, accurate echocardiographic measurement is a highly time-consuming and places a significant burden on clinical practice of echocardiography^3^.

The advent of deep learning models with computer vision has enabled fully automated image analysis and processing across a wide range of medical imaging^4–10^. Isolated task specific annotations in echocardiography have already been proposed in echocardiography^11,12^, including for both wall thickness^13^ and left ventricular ejection fraction^14,15^, however there are many additional measurements that have been tackled with deep learning models. Furthermore, commercially available models are not open-source, thus limiting the ability to build upon prior developments.

Automation through AI has the potential for more precision as well as more rapid turn-around^8,11,13,16–18^. In this study, we aimed to develop and validate a deep-learning framework (EchoNet-Measurements) for automated measurement of wide range of echocardiographic parameters and measurements. Using a pipeline of view classification and automated segmentation, EchoNet-Measurements was trained on the largest largest-scale echocardiography annotation dataset to enable automated comprehensive AI measurements.

## Methods

### Data Curation

For training and held-out test dataset, we identified 155,215 studies from 78,037 patients who received a clinical transthoracic echocardiogram at CSMC between April 2009 and June 2022 (**Figure 1**). A total of 18 key transthoracic echocardiographic (TTE) parameters were used for model training. Imaging data is saved in Digital Imaging and Communications in Medicine (DICOM) format and annotated by expert sonographers and cardiologists as part of routine practice.

**Figure 1:**
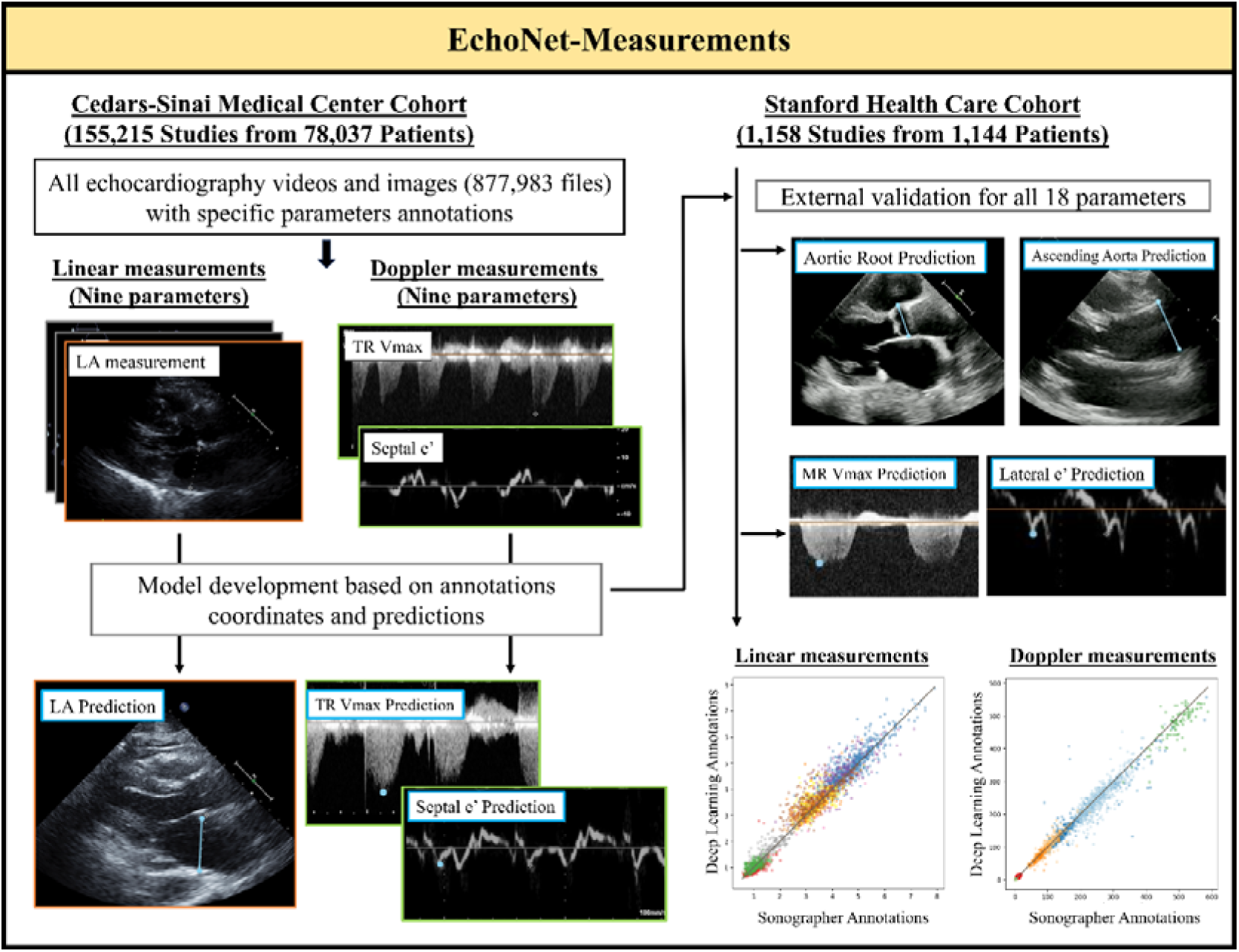
Overview of the study pipeline. The pipeline of the developed automated echocardiographic parameters measurement model includes two main groups: linear measurements (e.g., left atrium diameter and intraventricular septum) and Doppler measurements (e.g., tricuspid regurgitation peak velocity and septal e’ velocity). Evaluation of EchoNet-Measurements was performed on held-out test cohorts (CSMC) and external dataset (SHC), and the model demonstrated accuracy comparable to sonographer annotations. LA: left atrium; TR Vmax: tricuspid regurgitation maximum velocity.

Important linear measurements from B-mode echocardiographic images and videos included intraventricular septum (IVS), left ventricular internal diameter (LVID), left ventricular posterior wall (LVPW) diameter, left atrium diameter, right ventricular basal diameter, ascending aorta diameter, aortic root diameter, pulmonary artery diameter, and inferior vena cava (IVC). Important Doppler measurements include tricuspid regurgitation maximal velocity (TR Vmax), aortic valve maximal velocity (AV Vmax), mitral valve maximal velocity (MV Vmax), left ventricular outflow tract maximum velocity (LVOT Vmax) septal e’, and lateral e’, mitral valve peak E velocity, E/A, tricuspid annular plane systolic excursion (TAPSE).

Additionally, EchoNet-Measurements was evaluated on 1,158 studies from 1,144 patients from SHC who underwent echocardiography between January 2013 and August 2018 for external validation. Furthermore, to assess the end-to-end processing of EchoNet-Measurements for full TTE studies, 2,103 studies from 2,039 patients from June 2022 to November 2024 at CSMC were used as a temporal split validation dataset. All echocardiography studies in the CSMC development cohort dataset and the SHC cohort were performed using Philips EPIQ 7 or iE33 ultrasound machines. All echocardiography studies in the temporal split dataset at CSMC were conducted using Philips EPIQ 7 or EPIQ CVx ultrasound systems. Approval for this study was obtained from the Cedars-Sinai Medical Center and Stanford Healthcare Institutional Review Boards, and the requirement for informed consent was waived for retrospective data analysis without patient contact.

### Model Development

For EchoNet-Measurement training and testing, we used a DeepLabv3 architecture^19^ and trained with a binary cross-entropy with logits loss. The CSMC data was split by patient in a ratio of 8:1:1 for model training, validation, and held-out testing. Task specific models were trained for each measurement. For all models, an Adam optimizer with a learning rate of 0.001 was used, and the model was trained for up to 100 epochs with early stopping after 10 epochs based on the validation loss and a batch size of 24. The weights from the epoch with the minimum loss were used for the held-out test dataset and external validation. All performance metrics were analyzed using data from held-out data that was not involved in model training. An image quality classifier was also developed to identify low quality images and videos (examples in **Supplemental Figures 1**) which were excluded from downstream analysis. The image quality control model used a video-based convolutional neural network (R2+1D)^14^ to classify low-quality videos and an image-based model (DenseNet)^20^ to classify low-quality Doppler images^21^ or exclude inaccurate tissue doppler annotations (remove a’ annotation when e’ is desired). (**Supplemental Methods)**.

### Analysis of Cases with High Discrepancy between AI and clinicians

From the 90th percentile or higher of mean absolute error (MAE) between deep learning measurements and sonographer measurements, a total of 180 subsample data (randomly picked 10 images from each measurement variable) from the CSMC dataset were manually reviewed by two board-certified cardiologists (Y.S. and C.B.R). This analysis was conducted to identify the causes of significant differences between the deep-learning model measurements and those of the sonographers. We identified as if any of the following criteria applies: (1) the sonographer measurement (ground truth) was preferred over automatic measurements by the deep learning model, (2) the automatic measurements by the deep learning model were preferred over sonographer measurements, (3) images that both sonographer measurement and DL-model predictions are within a clinically-acceptable range but with a measurement variability, and (4) predictions on a severely low-quality images.

### Statistical analysis

MAE, coefficient of determination (R2) and intraclass correlation coefficients (ICC) between clinically measured parameters and parameters predicted by the developed deep-learning model were calculated in the held-out dataset and the external dataset. Bland-Altman plots where the average of the two measurements was plotted against the difference were used to check the agreement between the actual and predicted measurements. Stratified analyses were done by image-quality and patient characteristics including patient sex and a history of atrial fibrillation. All 95% confidence intervals were calculated with 10,000 bootstrapping samples. Data analysis was performed using both Python (version 3.10.12) and R (version 4.2.2) programming languages. This study was carried out following the CONSORT-AI guideline^22^.

### Code and Data Availability

The code, model weights, and demonstration video are available at https://github.com/echonet/measurement/. The patient data is not publicly available due to their potentially identifiable nature.

## RESULTS

### Model Development and Validation Cohorts

A total of 877,983 annotated measurements were identified from 155,215 CSMC echocardiography studies from 78,037 patients. The patient cohorts (mean age: 65.0 ± 17.4 years, female: 46.2% in CSMC derivation cohort) exhibited similar patient age, the proportion of female, race, the mean left ventricular ejection fraction and comorbidities across datasets. (**Table 1 and Supplementary Table 1-3)**. The number of training and validation examples for each parameter are summarized in **Table 2**.

**Table 1:** Participant characteristics.

|  | Derivation Cohort<br>(CSMC) | Temporal Split<br>Cohort (CSMC) | External Validation<br>Cohort (SHC) |
| --- | --- | --- | --- |
| Characteristic | N = 78,037<sup>1</sup> | N = 2,039<sup>1</sup> | N = 1,158<sup>1</sup> |
| Age, mean (SD) | 65.0 (17.4) | 66.2 (17.7) | 63.4 (18.8) |
| BMI, mean (SD) | 27.2 (6.4) | 26.4 (5.5) | 24.9 (9.0) |
| Gender, female | 35,829 (46.2%) | 921 (45.2%) | 567 (49.0) |
| Unknown | 555 | 1 | 15 |
| Race/Ethnicity |  |  |  |
| Asian | 5,638 (7.2%) | 201 (9.9%) | 193 (16.7) |
| Black or African<br>American | 10,593 (13.6%) | 279 (13.7%) | 65 (5.6) |
| Other | 8,427 (10.8%) | 260 (12.7%) | 264 (22.8) |
| White | 53,379 (68.4%) | 1,299 (63.7%) | 636 (54.9) |
| Hypertension | 38,122 (48.9%) | 794 (38.9%) | 581 (50.2) |
| Dyslipidemia | 36,779 (47.1%) | 883 (43.3%) | 575 (49.7) |
| Diabetes | 32,583 (41.8%) | 605 (29.7%) | 328 (28.3) |
| Atrial Fibrillation | 29,324 (37.6%) | 480 (23.5%) | 352 (30.4) |
| Heart Failure | 29,114 (37.3%) | 526 (25.8%) | 379 (32.7) |
| LVEF, mean (SD) | 58.0 (14.6) | 57.2 (13.0) | 58.1 (12.7) |
| Unknown | 4,585 |  |  |

<sup>1</sup>Mean (SD); n (%)
CSMC: Cedars-Sinai Medical Center, SHC: Stanford HealthCare, BMI: Body mass index, LVEF: Left ventricular ejection fraction. SD: Standard deviation

**Table 2:** Imaging characteristics.

|  | CSMC Training and Validation Cohorts |  |  |  | SHC External Validation Cohort |  |  |  |
| --- | --- | --- | --- | --- | --- | --- | --- | --- |
| Measurement | Number of Images | Number of Studies | Number of Patients | Mean measurement (SD) | Number of Images | Number of Studies | Number of Patients | Mean measurement (SD) |
| **Linear Measurements** |  |  |  |  |  |  |  |  |
| IVS | 97,685 | 94,311 | 55,284 | 1.13 (0.29) | 96 | 96 | 94 | 1.02 (0.21) |
| LVID | 224,042 | 131,873 | 71,321 | 3.71 (1.15) | 129 | 129 | 129 | 4.69 (0.78) |
| LVPW | 120,093 | 116,276 | 65,394 | 1.09 (0.25) | 55 | 55 | 54 | 1.01 (0.21) |
| Left Atrium | 89,779 | 89,170 | 56,805 | 3.87 (0.87) | 35 | 35 | 35 | 4.06 (0.72) |
| Ascending Aorta | 67,034 | 62,018 | 37,952 | 3.22 (0.54) | 77 | 77 | 77 | 3.5 (0.47) |
| Aortic root | 97,944 | 93,872 | 57,739 | 3.19 (0.73) | 32 | 32 | 32 | 3.25 (0.47) |
| RV Base | 92,729 | 90,389 | 53,566 | 3.68 (0.77) | 60 | 60 | 60 | 3.5 (0.6) |
| Pulmonary Artery | 8,960 | 6,875 | 5,723 | 2.2 (0.69) | 13 | 13 | 13 | 3.31 (0.78) |
| IVC | 32,288 | 31,815 | 21,171 | 1.68 (0.50) | 67 | 67 | 67 | 1.76 (0.5) |
| **Doppler Measurements** |  |  |  |  |  |  |  |  |
| TR Vmax | 199,243 | 79,897 | 47,584 | 248.6 (59.38) | 425 | 425 | 423 | 258.41 (46.07) |
| AV Vmax | 34,526 | 30,403 | 22,067 | 131.5 (41.86) | 320 | 320 | 320 | 170.62 (67.04) |
| MR Vmax | 16,819 | 12,417 | 10,301 | 411.0 (122.1) | 89 | 89 | 89 | 492.54 (50.08) |
| LVOT Vmax | 14,314 | 14,035 | 9,552 | 98.3 (32.39) | 304 | 304 | 304 | 101.22 (25.56) |
| Lateral e' | 96,051 | 95,578 | 58,495 | 9.33 (5.7) | 559 | 559 | 559 | 9.3 (3.32) |
| Septal e' | 43,462 | 43,343 | 33,327 | 7.07 (4.41) | 640 | 640 | 639 | 6.94 (2.51) |
| Peak E velocity | 62,698 | 62,331 | 43,060 | 82.8 (27.75) | 71 | 71 | 71 | 84.35 (26.73) |
| E/A | 62,698 | 62,331 | 43,060 | 1.24 (0.67) | 54 | 54 | 54 | 1.19 (0.79) |
| TAPSE | 54,344 | 53,802 | 36,189 | 1.95 (1.38) | 254 | 254 | 254 | 2.28 (0.5) |

SD: Standard deviation, IVS: Intraventricular septum, LVID: Left ventricular internal diameter, LVPW: Left ventricular posterior wall,
RV Base: Right ventricular basal diameter, IVC: Inferior vena cava, TR Vmax: Tricuspid regurgitation maximum velocity, AV Vmax:
Aortic valve maximum velocity, MR Vmax: Mitral regurgitation maximum velocity, LVOT Vmax: Left ventricular outflow tract
maximum velocity, Lateral e': Lateral mitral annulus e' velocity, Septal e': Septal mitral annulus e' velocity, Peak E velocity: Early
diastolic mitral inflow velocity, E/A: Ratio of early diastolic to late diastolic mitral inflow velocities. TAPSE: Tricuspid annular plane
systolic excursion.

### Assessment of EchoNet-Measurement’s Performance

In the CSMC test dataset, the EchoNet-Measurements model demonstrated a strong agreement between automated deep learning measurements and sonographer measurements with an overall R^2^ of 0.967 (0.965 – 0.970) in the held-out dataset. (ranging from a R^2^ = 0.27 (0.22-0.32) (ICC: 0.62 (0.60 – 0.63), MAE: 0.425 cm (0.404-0.446)) for ascending aorta diameter to R^2^ = 0.76 (ICC: 0.87 (0.86-0.87), MAE: 0.387cm (0.382-0.392)) for left ventricular internal diameter (LVID) (**Figure 2**)). Similarly for the Doppler images, the model demonstrated a high agreement between automated deep learning measurements and sonographer measurement, ranging from R^2^ = 0.55 (0.52-0.57) (ICC: 0.82 (0.66-0.89), MAE: 50.97 cm/2 (49.9-52.1)) for MR Vmax to R^2^ = 0.94 (ICC: 0.97 (0.97 – 0.97), MAE: 0.051 cm (0.050-0.052)) for TAPSE (**Table 3**).

**Figure 2:**
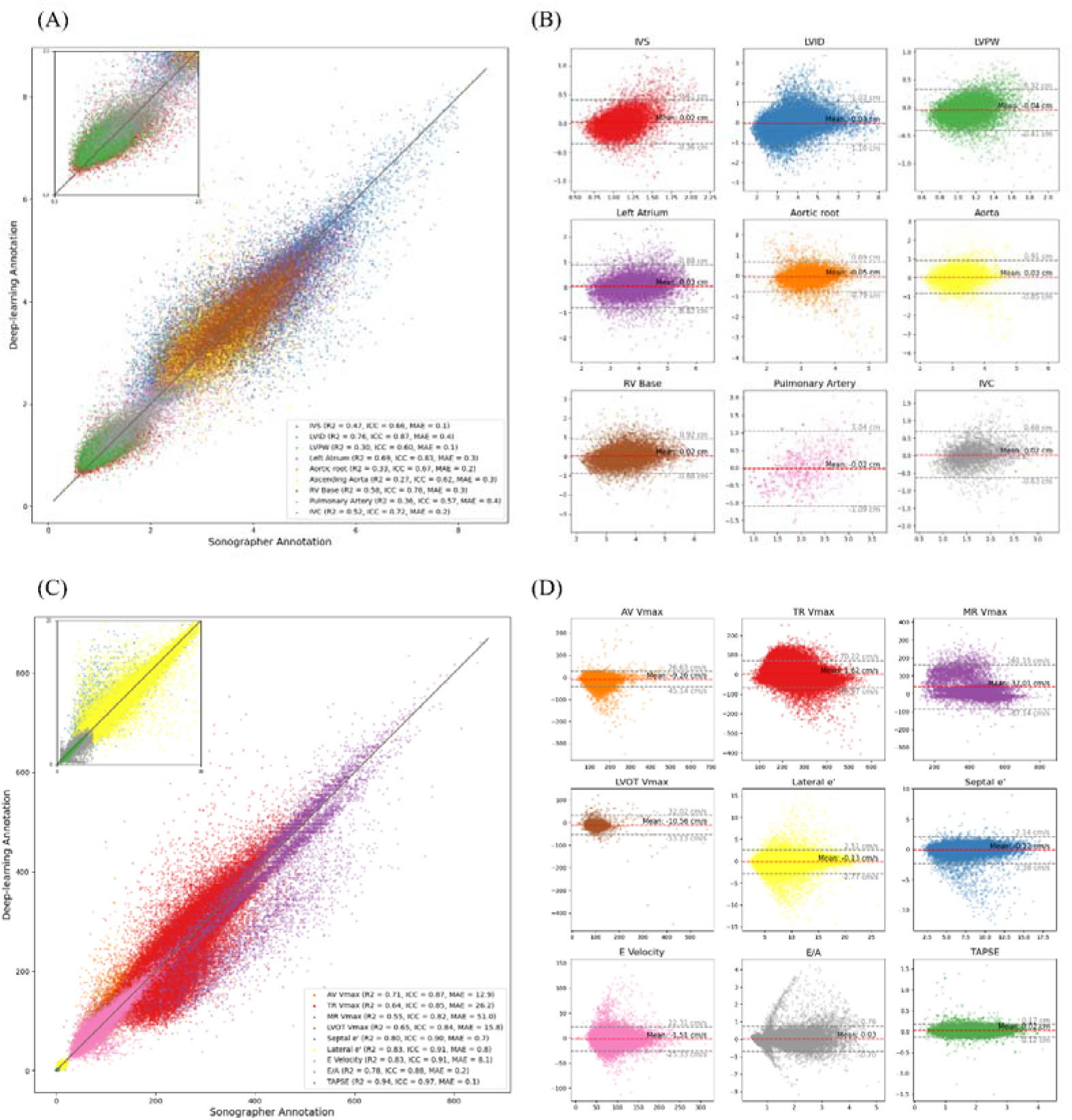
Model Performance and Agreement between Deep Learning Model and Sonographer Annotations for Echocardiographic Measurements in the CSMC test dataset. (A and C) Scatter plot comparing deep learning model predictions with sonographer annotations for nine linear echocardiographic parameters (A) and Doppler echocardiography parameters (C). Coefficient of determination (R²), intraclass correlation coefficient (ICC) and mean absolute error (MAE) are described in the legend. (B and D) Bland-Altman plots for each parameter in the linear measurement group (B) and Doppler echocardiography parameters (D), displaying the difference between model predictions and sonographer measurements (y-axis) against the mean of the two measurements (x-axis). Each plot includes the mean bias (red dashed line) and limits of agreement (±1.96 SD, gray dashed lines). For a detailed explanation of echocardiography parameter abbreviations, refer to Table 1.

**Table 3:** EchoNet-Measurements Performance in Internal and External Test Datasets.

|  | CSMC |  |  | SHC |  |  |
| --- | --- | --- | --- | --- | --- | --- |
|  | R<sup>2</sup> | ICC | MAE | R<sup>2</sup> | ICC | MAE |
| **Linear Measurements** |  |  |  |  |  |  |
| IVS | 0.468 (0.45-0.49) | 0.659 (0.64-0.67) | 0.143 (0.14-0.15) | 0.617 (0.46-0.72) | 0.78 (0.58-0.87) | 0.106 (0.09-0.12) |
| LVID | 0.765 (0.76-0.77) | 0.868 (0.86-0.87) | 0.386 (0.38-0.39) | 0.853 (0.76-0.91) | 0.922 (0.89-0.95) | 0.210 (0.17-0.25) |
| LVPW | 0.298 (0.27-0.32) | 0.602 (0.57-0.63) | 0.142 (0.14-0.14) | 0.712 (0.50-0.82) | 0.826 (0.72-0.89) | 0.093 (0.08-0.11) |
| Left Atrium | 0.689 (0.67-0.70) | 0.83 (0.82-0.84) | 0.314 (0.31-0.32) | 0.403 (-0.30-0.70) | 0.742 (0.47-0.87) | 0.410 (0.30-0.53) |
| Ascending Aorta | 0.271 (0.22-0.32) | 0.619 (0.6-0.63) | 0.308 (0.30-0.32) | 0.450 (0.08-0.66) | 0.757 (0.42-0.88) | 0.281 (0.24-0.33) |
| Aortic root | 0.332 (0.28-0.38) | 0.67 (0.66-0.68) | 0.245 (0.24-0.25) | 0.422 (-0.14-0.66) | 0.737 (0.24-0.9) | 0.286 (0.22-0.36) |
| RV Base | 0.585 (0.56-0.60) | 0.764 (0.76-0.77) | 0.340 (0.33-0.35) | 0.800 (0.64-0.90) | 0.896 (0.83-0.94) | 0.184 (0.14-0.23) |
| Pulmonary Artery | 0.361 (0.30-0.41) | 0.568 (0.53-0.61) | 0.425 (0.40-0.45) | 0.414 (-1.92-0.71) | 0.714 (0.11-0.91) | 0.489 (0.33-0.66) |
| IVC | 0.525 (0.49-0.56) | 0.723 (0.71-0.74) | 0.244 (0.24-0.25) | 0.738 (0.57-0.84) | 0.85 (0.62-0.93) | 0.185 (0.15-0.23) |
| **Doppler Measurements** |  |  |  |  |  |  |
| TR Vmax | 0.635 (0.63-0.64) | 0.846 (0.84-0.85) | 26.181 (26.05-26.32) | 0.786 (0.73-0.84) | 0.913 (0.9-0.93) | 15.392 (14.00-16.80) |
| AV Vmax | 0.707 (0.68-0.73) | 0.866 (0.77-0.91) | 12.853 (12.65-13.06) | 0.943 (0.91-0.97) | 0.977 (0.97-0.98) | 10.407 (9.14-11.84) |
| MR Vmax | 0.547 (0.52-0.57) | 0.824 (0.66-0.89) | 50.969 (49.85-52.13) | 0.323 (-0.14-0.60) | 0.796 (0.63-0.88) | 29.922 (24.45-36.16) |
| LVOT Vmax | 0.651 (0.51-0.75) | 0.844 (0.75-0.9) | 15.839 (15.23-16.49) | 0.808 (0.71-0.87) | 0.915 (0.88-0.94) | 7.886 (7.01-8.82) |
| Lateral e' | 0.835 (0.83-0.84) | 0.786 (0.71-0.84) | 0.834 (0.82-0.84) | 0.696 (0.62-0.76) | 0.845 (0.82-0.87) | 1.167 (1.05-1.29) |
| Septal e' | 0.801 (0.79-0.81) | 0.913 (0.91-0.91) | 0.709 (0.70-0.72) | 0.461 (0.33-0.57) | 0.735 (0.59-0.82) | 1.228 (1.12-1.33) |
| E velocity | 0.828 (0.82-0.84) | 0.915 (0.91-0.92) | 8.131 (8.02-8.24) | 0.810 (0.56-0.94) | 0.911 (0.74-0.96) | 6.796 (4.89-9.24) |
| E/A | 0.776 (0.76-0.79) | 0.882 (0.88-0.88) | 0.212 (0.21-0.22) | 0.365 (0.10-0.94) | 0.556 (0.34-0.72) | 0.215 (0.09-0.39) |
| TAPSE | 0.941 (0.87-0.98) | 0.97 (0.97-0.97) | 0.051 (0.05-0.05) | 0.512 (0.17-0.77) | 0.781 (0.72-0.83) | 0.148 (0.11-0.19) |

MAE: Mean absolute error. ICC: Intraclass Correlation Coefficient. For a detailed explanation of echocardiography parameter abbreviations, refer to Table 1. All metrics values are described with 95% confidence interval.

Representative images comparing sonographer annotations and deep learning annotations are shown in **Figure 3** and **Figure 4** for linear and Doppler measurements respectively. Representative generated videos in linear measurement group are shown in **Supplemental Video 1**. A demonstration video for the high-throughput analysis of TR Vmax using the input of multiple DICOM files is shown in **Supplemental Video 2**. Image quality greatly affected the performance of automated measurements, with the high-quality image group consistently showed a significant improvement in MAE and coefficient of determination compared to low-quality echocardiography images (**Supplementary Table 4 and Supplementary Figure 2-3**). The model performance was consistent across patient characteristics including sex and atrial fibrillation status in all measurement parameters (**Supplemental Table 5-6**)

**Figure 3:**
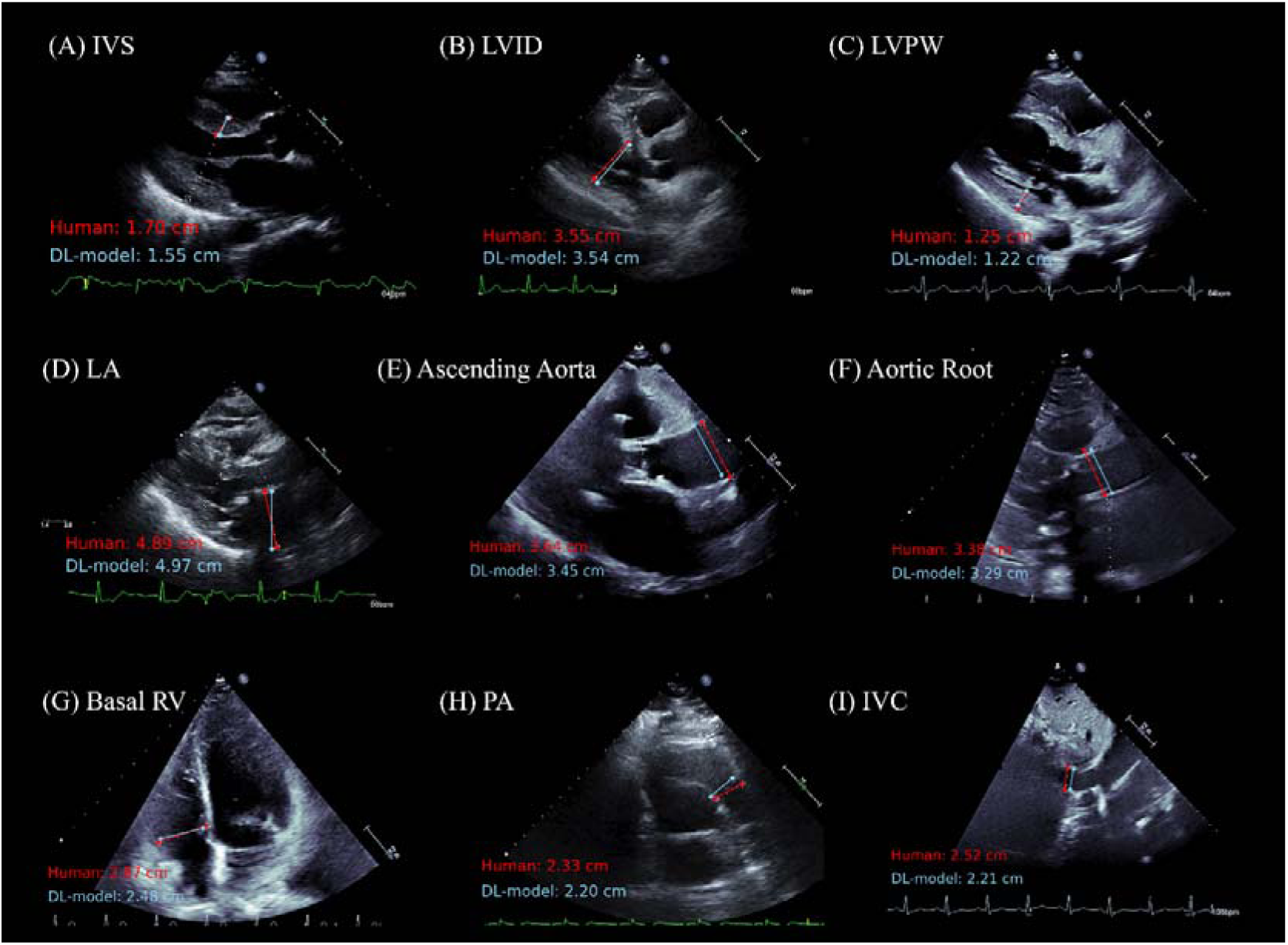
Representative Figures of Comparison between Sonographer and Deep Learning Model Measurements for Echocardiographic Linear Measurement Parameters. The figure showing a comparison of measurements made by a sonographer (in red) and predictions from the deep learning (DL) model (in light blue) across nine echocardiographic parameters. For a detailed explanation of echocardiography parameter abbreviations, refer to Table 1.

**Figure 4:**
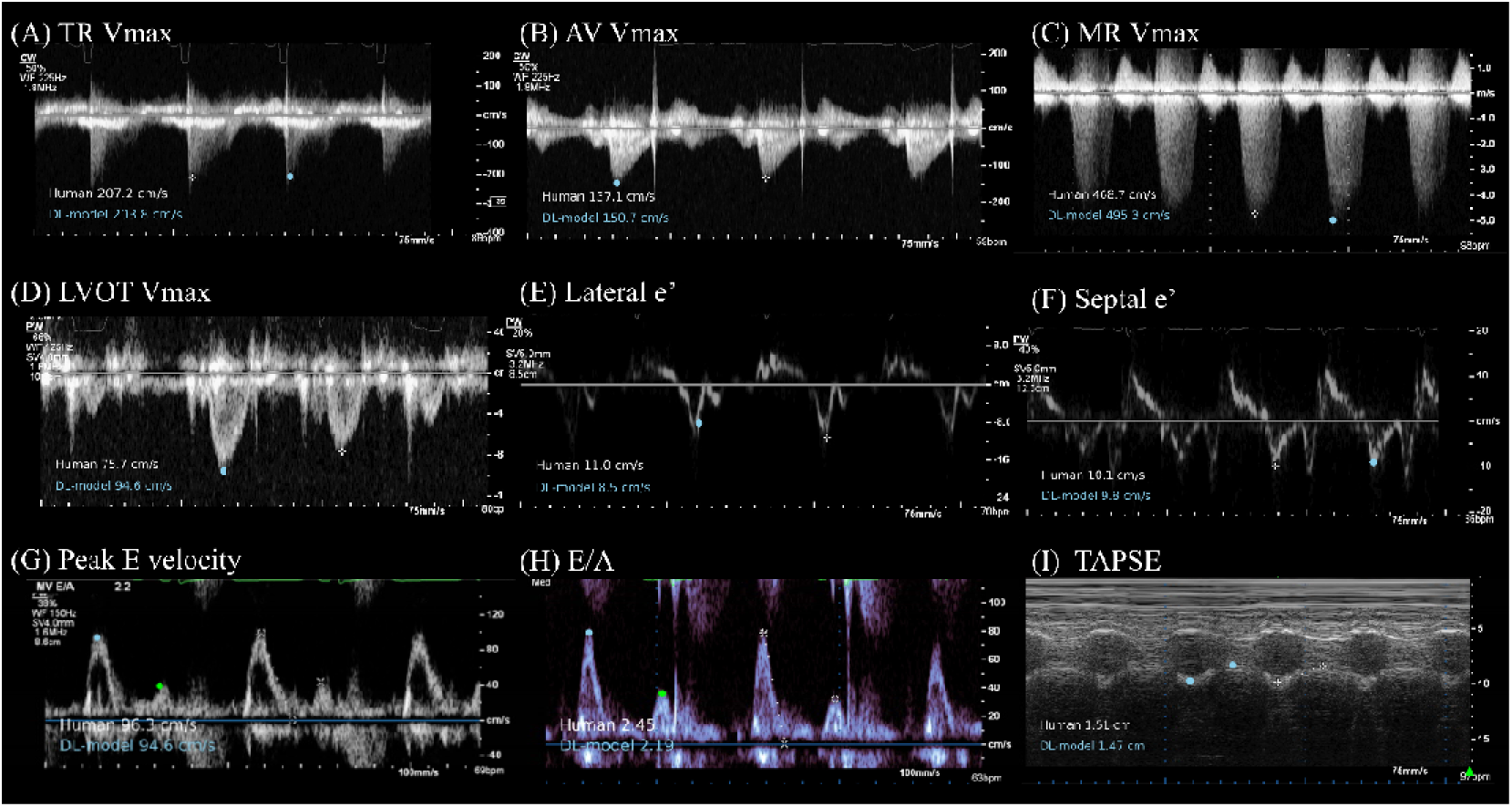
Representative Figures of Comparison between Sonographer and Deep Learning Model Measurements for Echocardiographic Doppler Measurement Parameters. Comparison of measurements made by a sonographer (in white) and predictions from the deep learning (DL) model (in light blue) across nine echocardiographic Doppler and M-mode parameters (TAPSE). For a detailed explanation of echocardiography parameter abbreviations, refer to Table 1. For Peak E velocity and E/A, deep-learning based annotation on peak E velocity is shown in light blue dot and peak A velocity is shown in green dot.

On the temporal split dataset from CSMC (June 2022 - November 2024), the model showed robust predictive accuracy (an overall R^2^ of 0.981 (0.976-0.984)) in end-to-end analysis (e.g., an R^2^ of 0.962 (0.95-0.97), an ICC of 0.981(0.98-0.98) and an MAE of 0.062 mm (95% CI 0.06-0.07) for TAPSE, and R^2^ of 0.748 (0.69-0.80), an ICC of 0.855 (0.77-0.90) and an MAE of 0.204mm (95% CI 0.19-0.22) for IVC diameter) (**Figure 5 and Supplemental Table 1)**.

**Figure 5:**
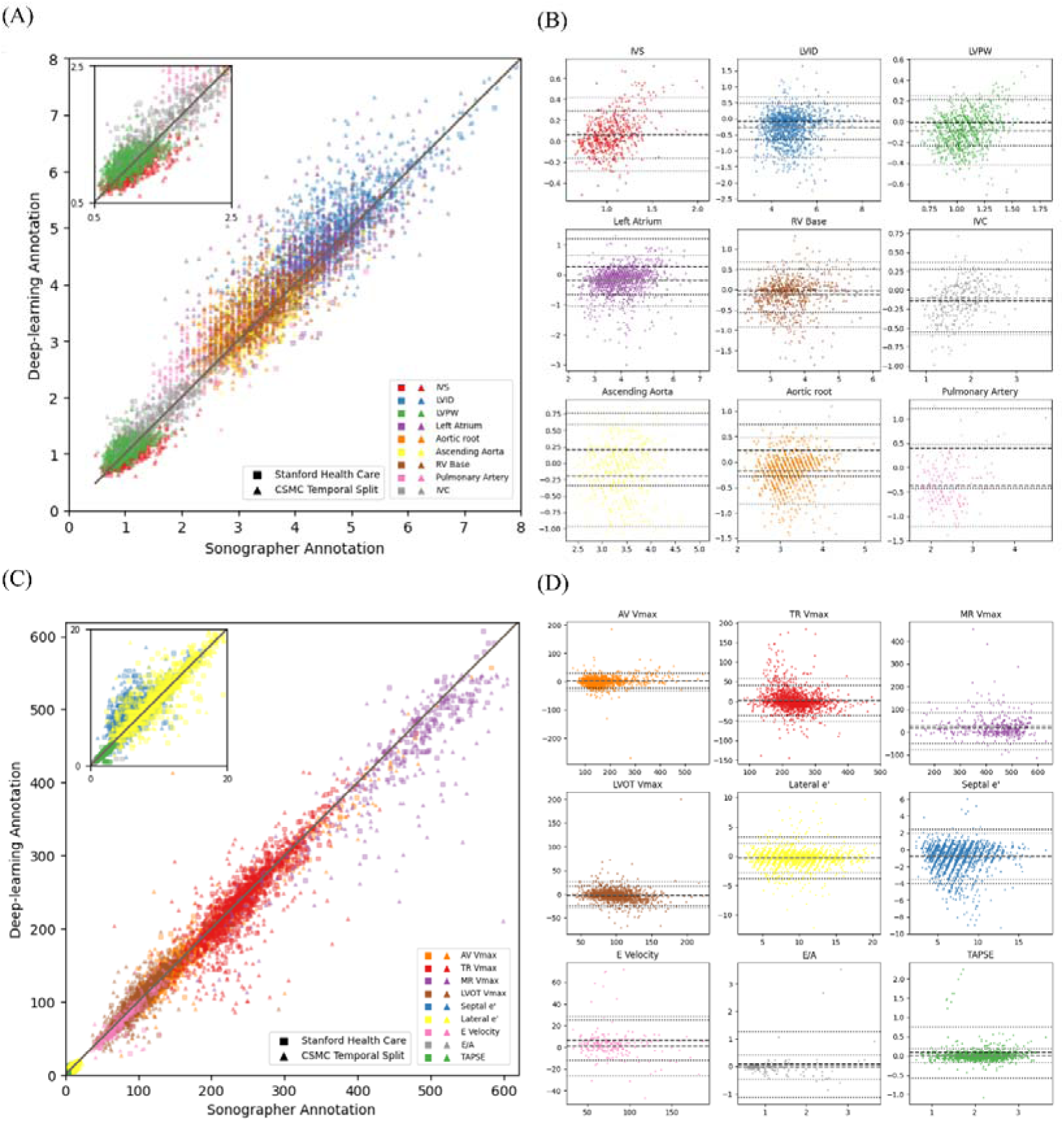
Model Performance and Agreement between Deep Learning Model and Sonographer Annotations for Echocardiographic Measurements in the SHC external test dataset and CSMC temporal split dataset. (A and C) Scatter plots comparing deep learning model predictions with sonographer annotations for (A) linear parameters and (C)Doppler echocardiography parameters. (B and D) Bland-Altman plots for each parameter in the linear measurement group (B) and Doppler echocardiography parameters (D). Data from SHC are described as square dots and in the CSMC temporal split data are shown in triangle dots in all figures. All metrics including coefficient of determination (R2), intraclass correlation coefficients (ICC), mean absolute error (MAE), bias and limits of agreement are described in **Supplemental Tables 1 and 2**.

In the external SHC dataset with image-level ground truth measurement value, EchoNet-Measurement’s performance was robust (**Figure 5 and Supplemental Table 2)** with an overall R^2^ of 0.987 (0.984 – 0.989) in the SHC dataset. The average R^2^ for linear measurements was 0.961 (0.95-0.97) with MAE ranging from 0.106mm (0.09-0.12) for IVS diameter to 0.489mm (0.34-0.66) for pulmonary artery diameter. The model’s performance was also consistent and robust across all Doppler parameters with an overall R^2^ of 0.990 (0.988 – 0.991) for linear measurements (e.g., MAE of 0.148 cm (0.11-0.19) in TAPSE and 7.89 mmHg (7.01-8.82) in LVOT Vmax). The model performance was similarly robust in external dataset with study-level ground truth label for both linear measurement and all Doppler parameters with an overall R^2^ of 0.942 (0.936-0.947) and 0.983 (0.979-0.986), respectively (**Supplemental Table 3, Supplemental Figure 4**)

### Analysis of High Measurement Discrepancies Cases

Among echocardiography images with absolute differences between sonographer measurement and deep-learning measurement in the 90th percentile or higher, a subset of 180 images (randomly selected 10 images from each measurement variable) were manually reviewed. Of a total of 180 images, 91 (50.6%) were categorized as the cases where sonographer annotations were preferred over automatic measurements by the deep learning model, 20 images (11.1%) was classified as the images where the deep-learning model was preferred over sonographer measurements, and 54 images (30.0%) were categorized the images that both sonographer annotations and deep-learning model predictions were clinically acceptable but demonstrating a measurement variabilities. In the remaining 15 images (8.3%), echocardiography images were severely noisy or blurred, seen as inappropriate images for deep learning-based measurements. Representative images are described in **Supplemental Figure 5** and the number of images in each category is described in **Supplemental Table 7**.

## DISCUSSION

In this study, we developed EchoNet-Measurements, a deep learning model for the automatic measurement of 18 echocardiographic parameters, and showed the model performed well in multiple geographically and temporally distinct cohorts. Using the largest yet real-world dataset of sonographer clinical annotations, this open-source model demonstrated favorable measurement accuracy comparable to that of expert sonographers. Further, we have released the code and weights of these models publicly, along with demo user interface, to facilitate further research in the field.

Comprehensive measurement is needed since clinical echocardiography involves the measurement of many parameters that holistically assess patient status. For example, diagnosis for heart failure with preserved ejection fraction (HFpEF) requires a combination of multiple echocardiographic parameters^23,24^—including left atrial volume index (LAVI), lateral e’, septal e’, E/A ratio, right ventricular systolic pressure), and TR velocity. Additionally, measuring the IVC diameter and its collapsibility is useful for estimating left ventricular filling pressures and right atrial pressure, which are critical for the serial surveillance of heart failure status.^25,26^ One of the primary benefits of measurement automation is the reduction of examination time and variability between sonographers, leading to more consistent result.^27,28^

Our study has both strengths and limitations. Previous studies have already focused on automating the measurement of limited echocardiographic parameters using deep learning segmentation models^11,16,17^, but we note that many prior models do not share code or model weights - limiting utility for reproducible research purposes and future development. EchoNet-Measurements is trained on data from only one medical center, which might capture biases and quirks associated with the dataset, however we demonstrate favorable results compared to previous deep-learning models^13,17,18,29^. Furthermore, one of the strengths of our model is the size of the training data, which is the largest yet dataset of sonographer annotations. In multiple prior works both within medicine^30^ and outside^31^, the scale of training data has a strong impact on downstream model performance. Another features of our work is the public availability of our model and demo interface, which enables the creation of clinical datasets and supports reproducible clinical research.

## Conclusion

In this study, we developed and validated EchoNet-Measurements, an open-source deep learning framework for the automated assessment of a wide range of echocardiographic parameters. Trained on the largest dataset to date of clinical annotations, the model demonstrated robust performance across both linear and Doppler measurements.

## Data Availability

Code and Data Availability
The code, model weights, and demonstration video are available at https://github.com/echonet/measurement/. The patient data is not publicly available due to their potentially identifiable nature.

https://github.com/echonet/measurement/

## Abbreviations and Acronyms

CSMC: Cedars-Sinai Medical Center
SHC: Stanford Healthcare
DICOM: Digital Imaging and Communications in Medicine
LVOT Vmax: Left Ventricular Outflow Tract Maximum Velocity
TR Vmax: Tricuspid Regurgitation Maximum Velocity
IVS: Intraventricular Septum
LVID: Left Ventricular Internal Diameter
LVPW: Left Ventricular Posterior Wall
TAPSE: Tricuspid Annular Plane Systolic Excursion
MAE: Mean Absolute Error
AV Vmax: Aortic Valve Maximum Velocity
MV Vmax: Mitral Valve Maximum Velocity
E/A: Ratio of Early (E) to Late (A) Diastolic Mitral Flow Velocities
IVC: Inferior Vena Cava

## Supplemental Materials

**Supplemental Table 1:** Dataset characteristics and model Performance in temporal split CSMC dataset (June 2022 – November 2024)

| Variable | total file | unique study | unique patient | Mean Sonographer<br>Measurement (SD) | R<sup>2</sup> (95% CI) | ICC (95% CI) | MAE (95% CI) | Bias and limits of<br>agreement |
| --- | --- | --- | --- | --- | --- | --- | --- | --- |
| **Linear Measurements** |  |  |  |  |  |  |  |  |
| IVS | 539 | 539 | 533 | 1.09 (0.26) | 0.471 (0.39-0.53) | 0.665 (0.56-0.74) | 0.138 (0.13-0.15) | 0.07: -0.28 to 0.42 cm |
| LVID | 1,153 | 1,153 | 1,132 | 4.69 (0.81) | 0.529 (0.47-0.58) | 0.765 (0.57-0.86) | 0.403 (0.38-0.42) | -0.28: -1.23 to 0.66 cm |
| LVPW | 837 | 837 | 824 | 1.04 (0.22) | 0.207 (0.10-0.30) | 0.56 (0.38-0.68) | 0.151 (0.14-0.16) | -0.09: -0.42 to 0.25 cm |
| Left Atrium | 1,252 | 1,252 | 1,229 | 3.89 (0.82) | 0.659 (0.60-0.71) | 0.818 (0.71-0.88) | 0.322 (0.31-0.34) | -0.20: -1.05 to 0.64 cm |
| Ascending Aorta | 303 | 303 | 301 | 3.29 (0.47) | 0.144<br>(-0.00-0.30) | 0.594 (0.42-0.71) | 0.341 (0.30-0.37) | -0.19: -0.96 to 0.59 cm |
| Aortic root | 799 | 799 | 785 | 3.13 (0.47) | 0.351 (0.23-0.44) | 0.674 (0.49-0.78) | 0.273 (0.25-0.29) | -0.18: -0.83 to 0.48 cm |
| RV Base | 710 | 710 | 701 | 3.52 (0.7) | 0.633 (0.55-0.70) | 0.806 (0.76-0.84) | 0.292 (0.27-0.32) | -0.12: -0.92 to 0.68 cm |
| Pulmonary Artery | 142 | 142 | 138 | 2.19 (0.48) | -0.411<br>(-1.12-0.01) | 0.447 (0.06-0.67) | 0.469 (0.42-0.52) | -0.37: -1.20 to 0.47 cm |
| IVC | 269 | 269 | 263 | 1.76 (0.53) | 0.748 (0.69-0.80) | 0.855 (0.77-0.9) | 0.204 (0.19-0.22) | -0.11: -0.59 to 0.37 cm |
| **Doppler Measurements** |  |  |  |  |  |  |  |  |
| TR Vmax | 1,343 | 1,343 | 1,310 | 234.15 (53.03) | 0.715 (0.66-0.76) | 0.871 (0.86-0.88) | 16.289<br>(15.37-17.19) | 3.47: -51.57 to 58.51 cm/s |
| AV Vmax | 1,234 | 1,234 | 1,212 | 163.31 (68.45) | 0.941 (0.91-0.96) | 0.97 (0.97-0.97) | 9.165 (8.52-9.97) | 0.96: -31.54 to 33.46 cm/s |
| MR Vmax | 358 | 358 | 350 | 433.66 (101.74) | 0.662 (0.52-0.79) | 0.839 (0.73-0.9) | 33.523<br>(29.32-38.38) | 26.22: -77.55 to 130.00<br>cm/s |
| LVOT Vmax | 961 | 961 | 947 | 99.34 (24.88) | 0.671 (0.54-0.75) | 0.851 (0.83-0.87) | 8.711 (8.15-9.48) | -1.00: -28.88 to 26.88 cm/s |
| Lateral e' | 1,364 | 1,364 | 1,340 | 9.39 (3.46) | 0.854 (0.82-0.88) | 0.927 (0.91-0.94) | 0.828 (0.79-0.88) | -0.29: -2.82 to 2.25 cm/s |
| Septal e' | 1,330 | 1,330 | 1,314 | 6.99 (2.52) | 0.593 (0.52-0.65) | 0.808 (0.66-0.88) | 1.048 (0.99-1.10) | -0.77: -3.53 to 1.99 cm/s |
| Peak E velocity | 168 | 168 | 168 | 81.66 (28.93) | 0.765 (0.59-0.89) | 0.893 (0.86-0.92) | 7.806 (6.14-9.71) | 1.07: -26.24 to 28.38 cm/s |
| E/A | 108 | 108 | 108 | 1.25 (0.64) | 0.868 (0.74-0.92) | 0.936 (0.91-0.96) | 0.140 (0.11-0.18) | -0.03: -0.48 to 0.42 |
| TAPSE | 981 | 981 | 975 | 2.09 (0.48) | 0.962 (0.95-0.97) | 0.981 (0.98-0.98) | 0.062 (0.06-0.07) | 0.00: -0.19 to 0.19 cm |

IVS: Intraventricular septum, LVID: Left ventricular internal diameter, LVPW: Left ventricular posterior wall, RV Base: Right ventricular basal diameter, IVC: Inferior vena cava, TR Vmax: Tricuspid regurgitation maximum velocity, AV Vmax: Aortic valve maximum velocity, MR Vmax: Mitral regurgitation maximum 0 velocity, LVOT Vmax: Left ventricular outflow tract maximum velocity, Lateral e': Lateral mitral annulus e' velocity, Septal e': Septal mitral annulus e' velocity, TAPSE: Tricuspid annular plane systolic excursion, CSMC: Cedars-Sinai Medical Center

**Supplemental Table 2:**
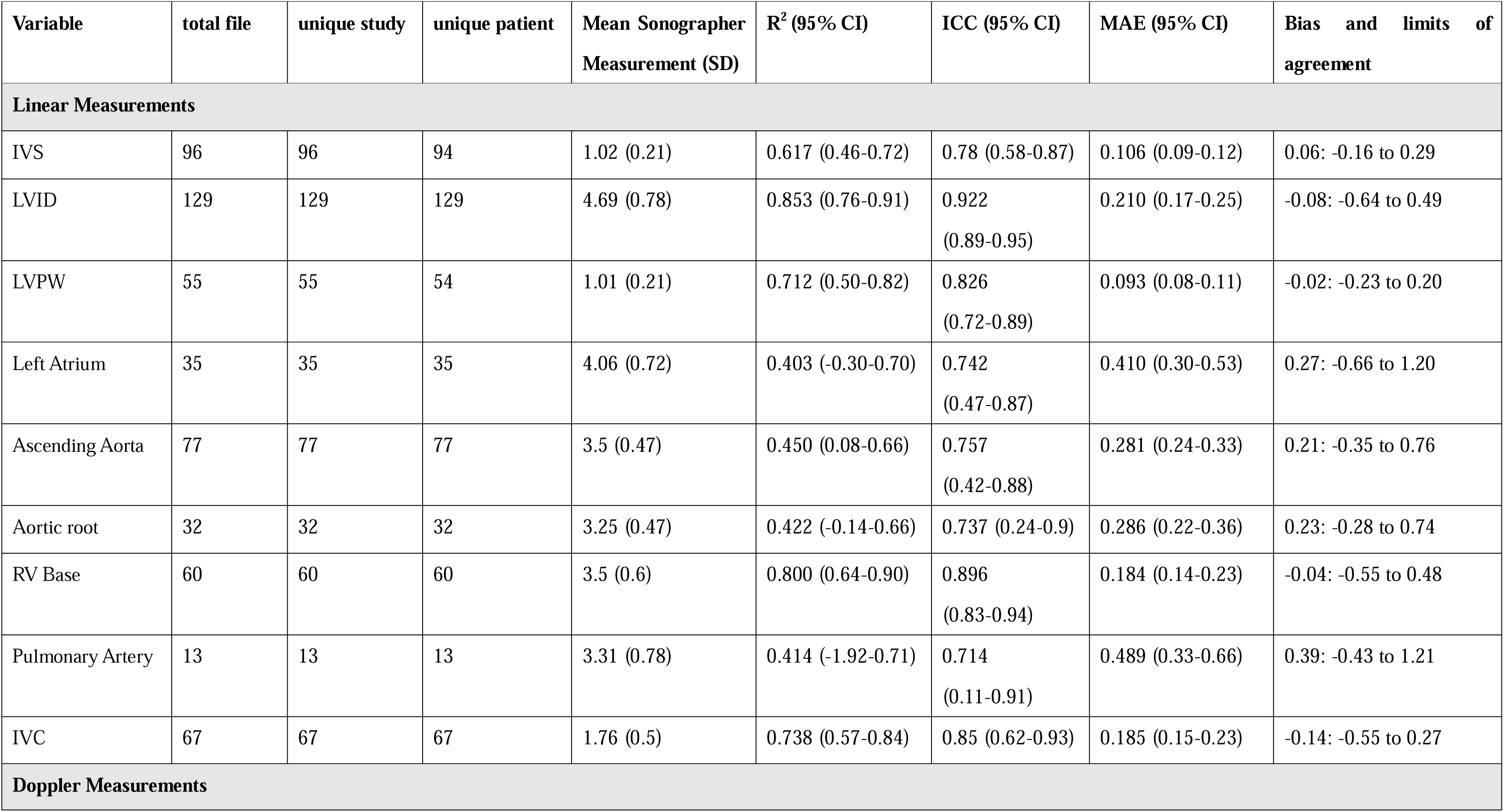
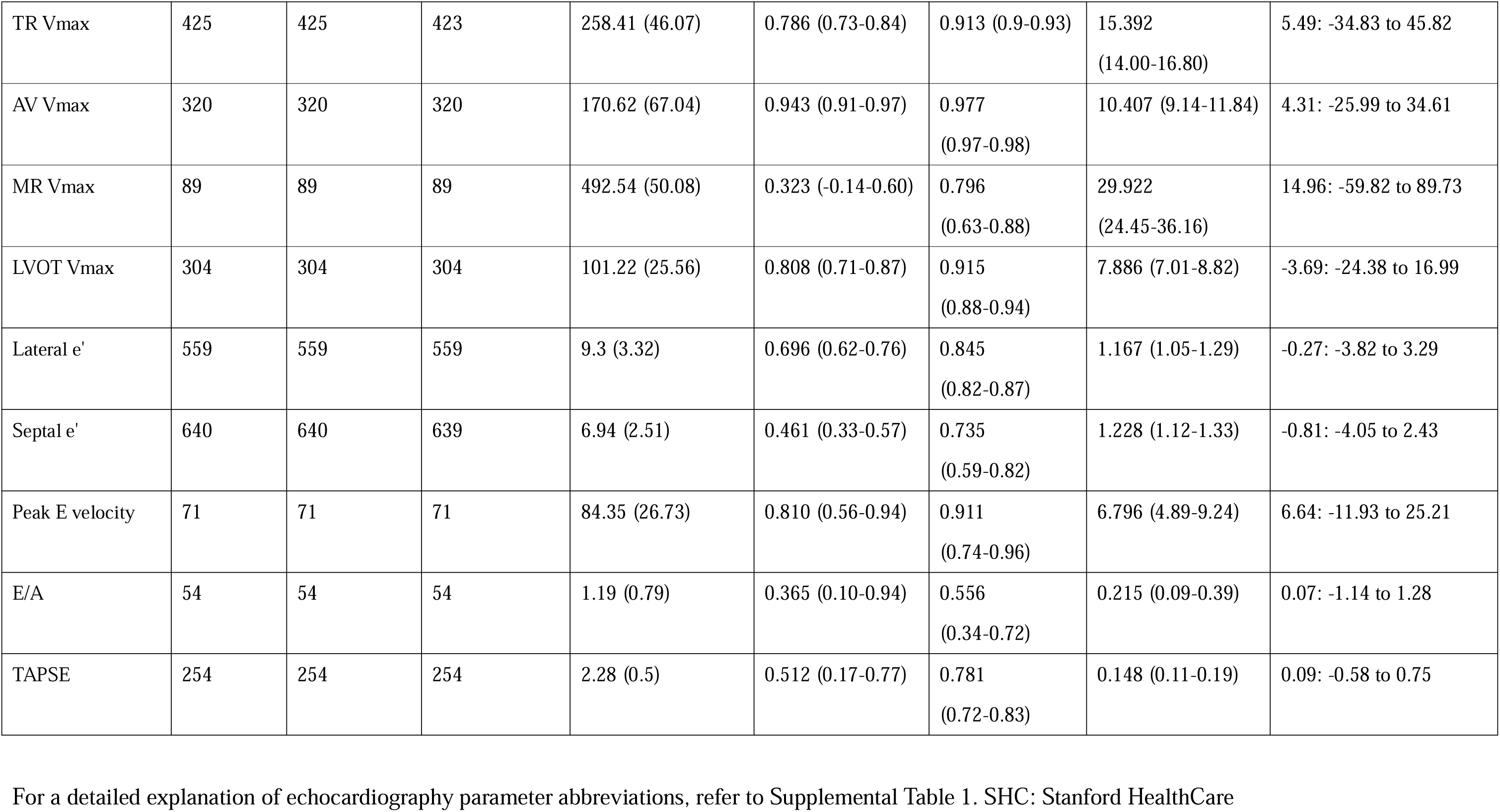
External dataset characteristics and model Performance in Stanford Health Care (Dataset with image-level ground truth)

| Variable | total file | unique study | unique patient | Mean Sonographer Measurement (SD) | R<sup>2</sup> (95% CI) | ICC (95% CI) | MAE (95% CI) | Bias and limits of agreement |
| --- | --- | --- | --- | --- | --- | --- | --- | --- |
| **Linear Measurements** |  |  |  |  |  |  |  |  |
| IVS | 96 | 96 | 94 | 1.02 (0.21) | 0.617 (0.46-0.72) | 0.78 (0.58-0.87) | 0.106 (0.09-0.12) | 0.06: -0.16 to 0.29 |
| LVID | 129 | 129 | 129 | 4.69 (0.78) | 0.853 (0.76-0.91) | 0.922 (0.89-0.95) | 0.210 (0.17-0.25) | -0.08: -0.64 to 0.49 |
| LVPW | 55 | 55 | 54 | 1.01 (0.21) | 0.712 (0.50-0.82) | 0.826 (0.72-0.89) | 0.093 (0.08-0.11) | -0.02: -0.23 to 0.20 |
| Left Atrium | 35 | 35 | 35 | 4.06 (0.72) | 0.403 (-0.30-0.70) | 0.742 (0.47-0.87) | 0.410 (0.30-0.53) | 0.27: -0.66 to 1.20 |
| Ascending Aorta | 77 | 77 | 77 | 3.5 (0.47) | 0.450 (0.08-0.66) | 0.757 (0.42-0.88) | 0.281 (0.24-0.33) | 0.21: -0.35 to 0.76 |
| Aortic root | 32 | 32 | 32 | 3.25 (0.47) | 0.422 (-0.14-0.66) | 0.737 (0.24-0.9) | 0.286 (0.22-0.36) | 0.23: -0.28 to 0.74 |
| RV Base | 60 | 60 | 60 | 3.5 (0.6) | 0.800 (0.64-0.90) | 0.896 (0.83-0.94) | 0.184 (0.14-0.23) | -0.04: -0.55 to 0.48 |
| Pulmonary Artery | 13 | 13 | 13 | 3.31 (0.78) | 0.414 (-1.92-0.71) | 0.714 (0.11-0.91) | 0.489 (0.33-0.66) | 0.39: -0.43 to 1.21 |
| IVC | 67 | 67 | 67 | 1.76 (0.5) | 0.738 (0.57-0.84) | 0.85 (0.62-0.93) | 0.185 (0.15-0.23) | -0.14: -0.55 to 0.27 |
| **Doppler Measurements** |  |  |  |  |  |  |  |  |
| TR Vmax | 425 | 425 | 423 | 258.41 (46.07) | 0.786 (0.73-0.84) | 0.913 (0.9-0.93) | 15.392<br>(14.00-16.80) | 5.49: -34.83 to 45.82 |
| AV Vmax | 320 | 320 | 320 | 170.62 (67.04) | 0.943 (0.91-0.97) | 0.977<br>(0.97-0.98) | 10.407 (9.14-11.84) | 4.31: -25.99 to 34.61 |
| MR Vmax | 89 | 89 | 89 | 492.54 (50.08) | 0.323 (-0.14-0.60) | 0.796<br>(0.63-0.88) | 29.922<br>(24.45-36.16) | 14.96: -59.82 to 89.73 |
| LVOT Vmax | 304 | 304 | 304 | 101.22 (25.56) | 0.808 (0.71-0.87) | 0.915<br>(0.88-0.94) | 7.886 (7.01-8.82) | -3.69: -24.38 to 16.99 |
| Lateral e' | 559 | 559 | 559 | 9.3 (3.32) | 0.696 (0.62-0.76) | 0.845<br>(0.82-0.87) | 1.167 (1.05-1.29) | -0.27: -3.82 to 3.29 |
| Septal e' | 640 | 640 | 639 | 6.94 (2.51) | 0.461 (0.33-0.57) | 0.735<br>(0.59-0.82) | 1.228 (1.12-1.33) | -0.81: -4.05 to 2.43 |
| Peak E velocity | 71 | 71 | 71 | 84.35 (26.73) | 0.810 (0.56-0.94) | 0.911<br>(0.74-0.96) | 6.796 (4.89-9.24) | 6.64: -11.93 to 25.21 |
| E/A | 54 | 54 | 54 | 1.19 (0.79) | 0.365 (0.10-0.94) | 0.556<br>(0.34-0.72) | 0.215 (0.09-0.39) | 0.07: -1.14 to 1.28 |
| TAPSE | 254 | 254 | 254 | 2.28 (0.5) | 0.512 (0.17-0.77) | 0.781<br>(0.72-0.83) | 0.148 (0.11-0.19) | 0.09: -0.58 to 0.75 |

For a detailed explanation of echocardiography parameter abbreviations, refer to Supplemental Table 1. SHC: Stanford HealthCare

**Supplemental Table 3:**
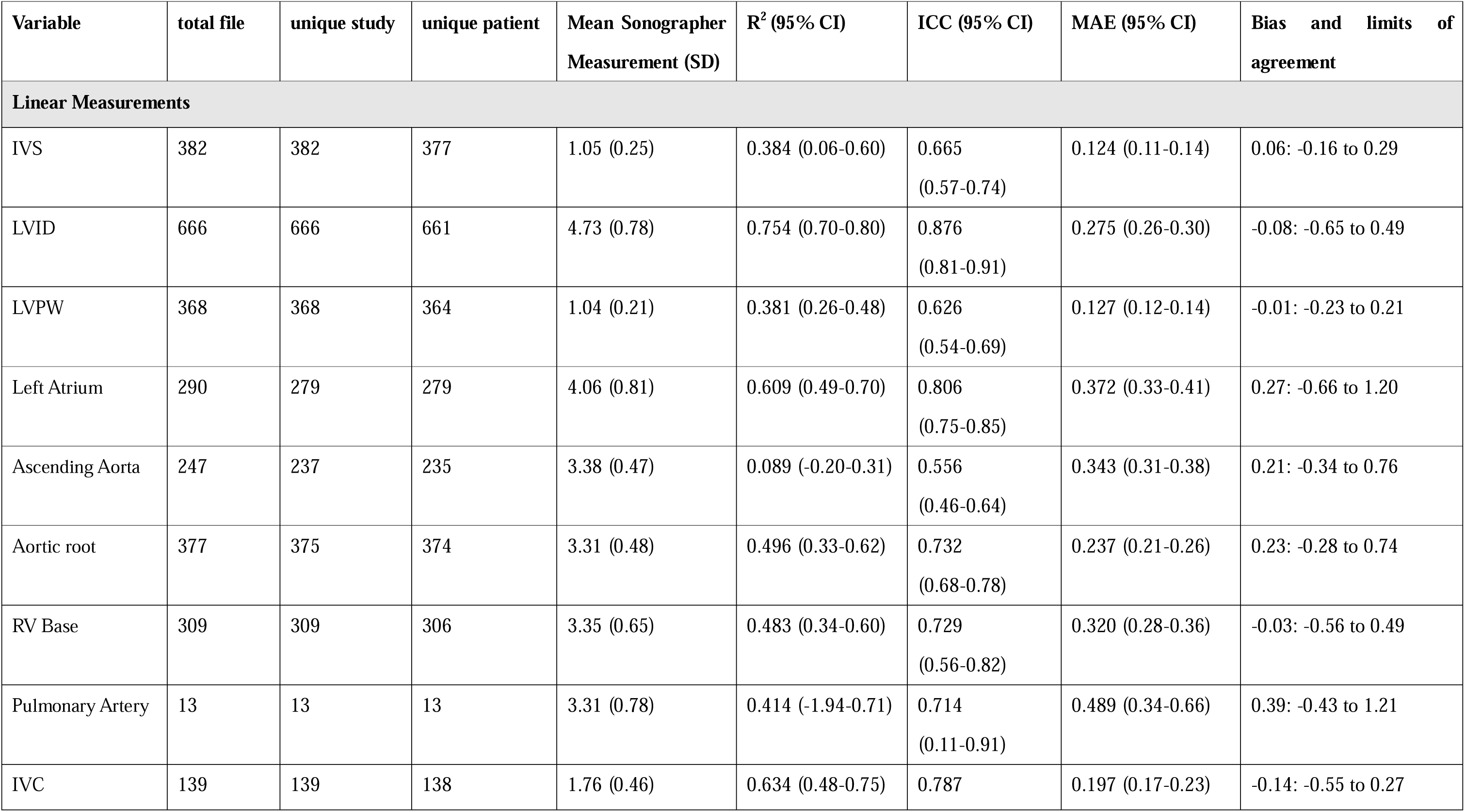
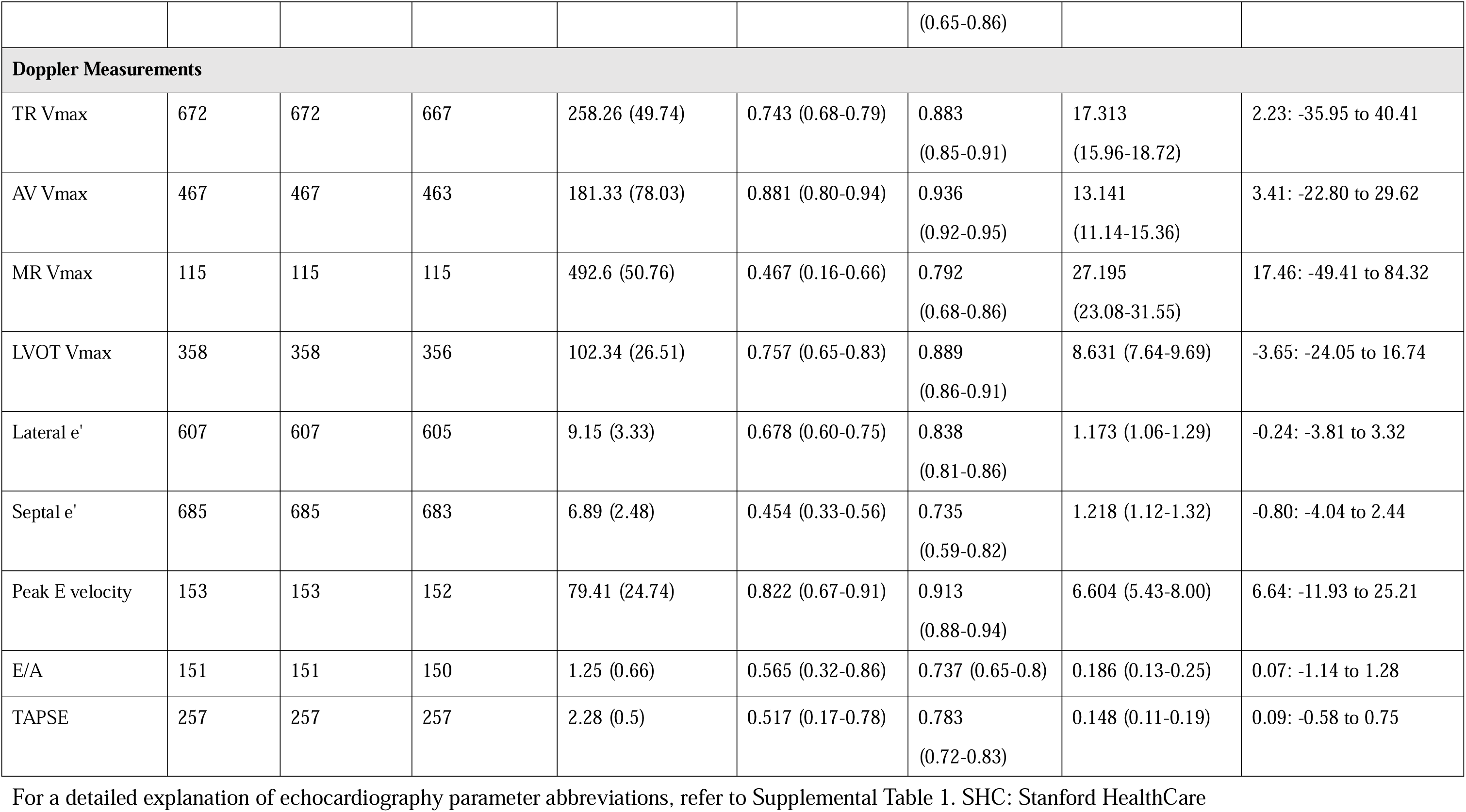
External dataset characteristics and model Performance in Stanford Health Care (Dataset with study-level ground truth cohort)

**Supplemental Table 4:**
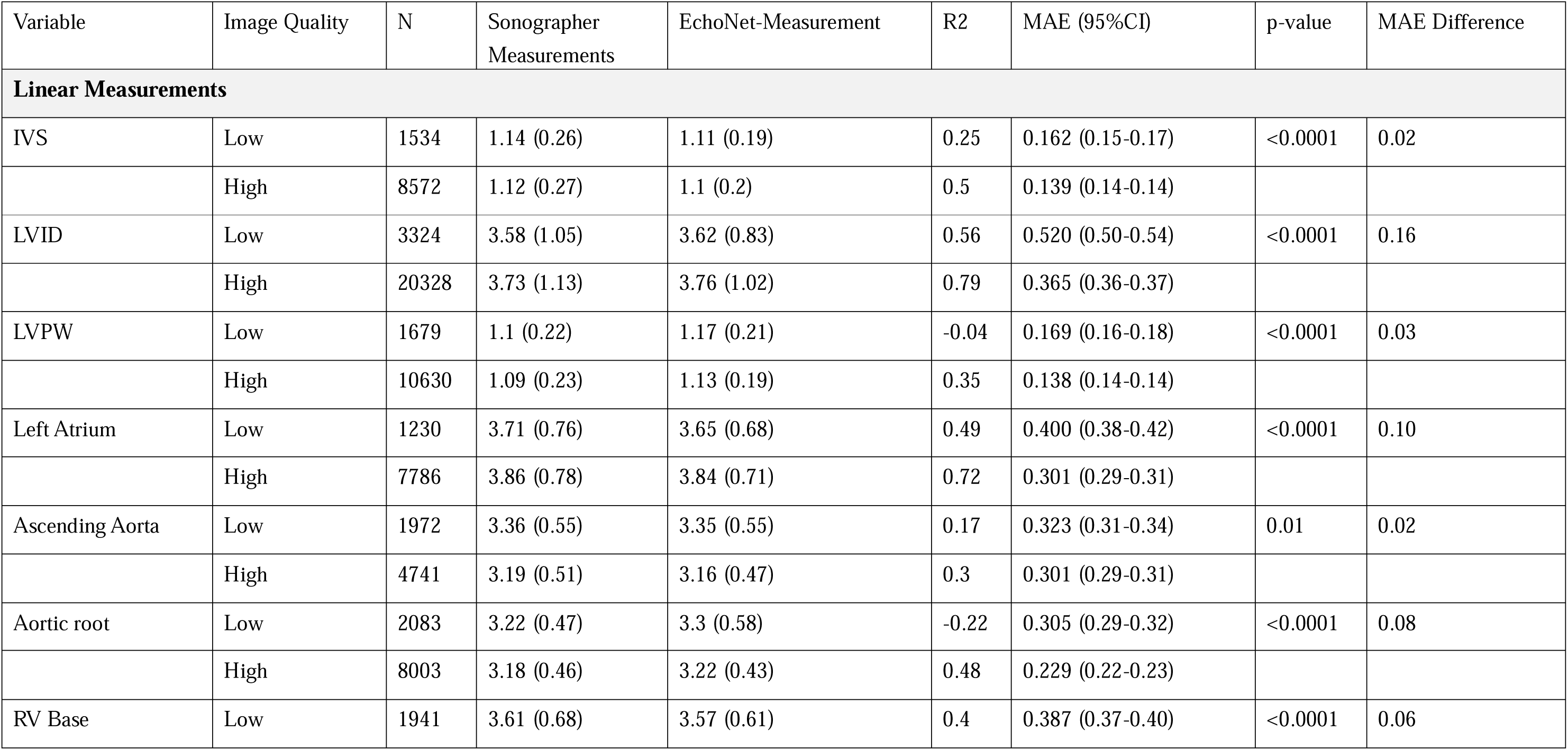
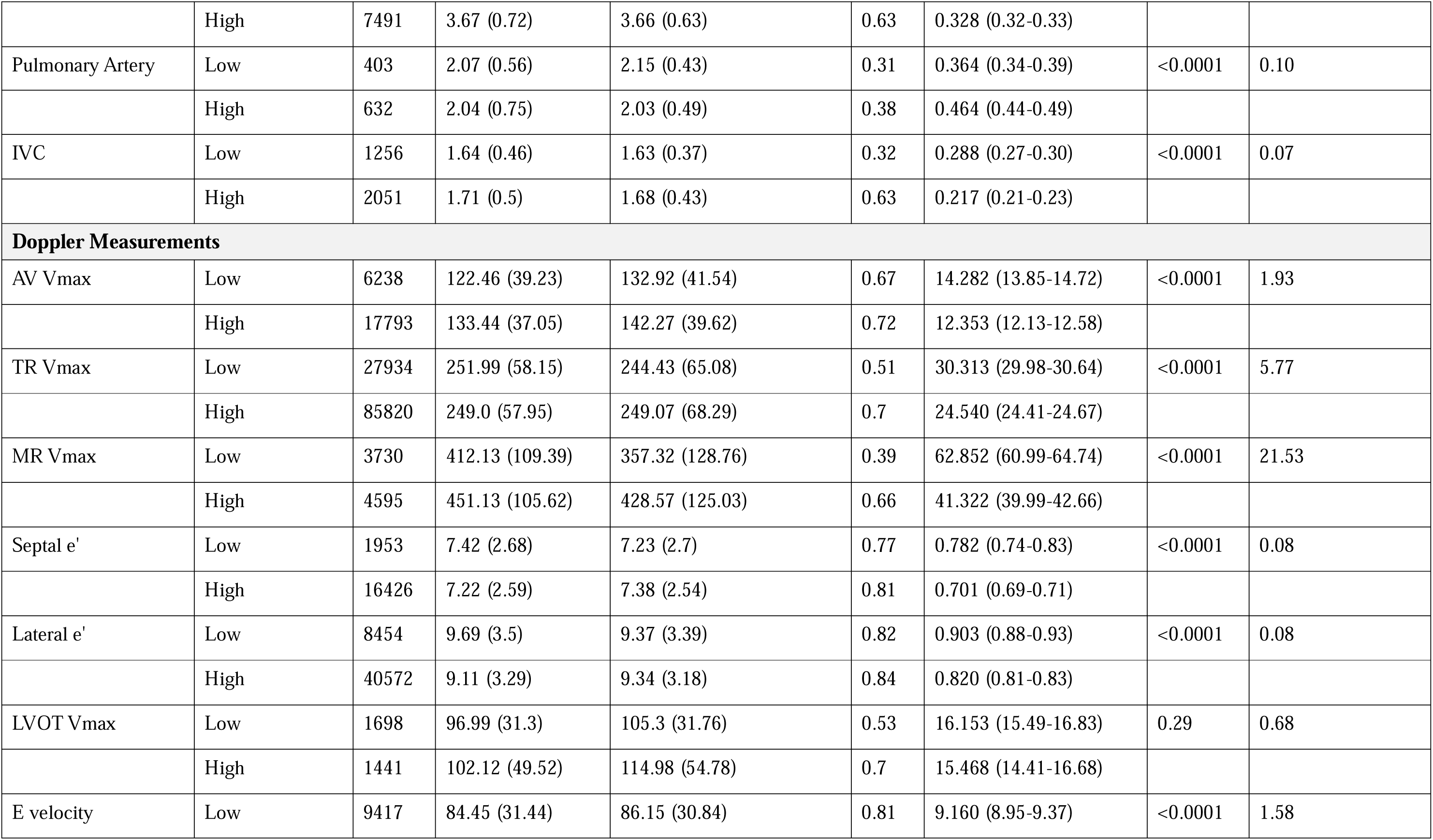
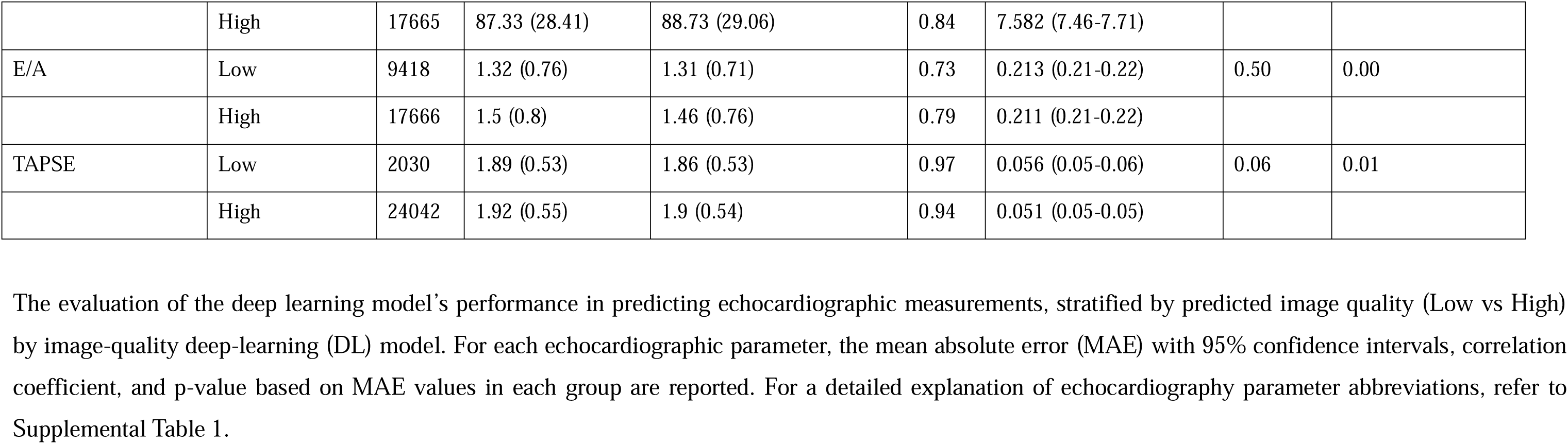
Evaluation of Deep Learning Model Performance by Predicted Image Quality for Echocardiographic Parameters (CSMC Held-Out Dataset)

**Supplemental Table 5:** Evaluation of Deep Learning Model Performance by Patient Sex (CSMC Held-Out Dataset)

| Variable | Sex | N | Sonographer<br>Measurements | DL-model<br>Measurements | R2 | MAE (95%CI) | p-value | MAE Difference |
| --- | --- | --- | --- | --- | --- | --- | --- | --- |
| **Linear Measurements** |  |  |  |  |  |  |  |  |
| IVS | Male | 5778 | 1.16 (0.26) | 1.14 (0.19) | 0.44 | 0.145 (0.14-0.15) | 0.06 | 0.005 |
|  | Female | 4328 | 1.08 (0.27) | 1.05 (0.19) | 0.48 | 0.140 (0.14-0.14) |  |  |
| LVID | Male | 13419 | 3.92 (1.15) | 3.95 (1.02) | 0.76 | 0.397 (0.39-0.40) | <0.0001 | 0.025 |
|  | Female | 10233 | 3.43 (1.02) | 3.46 (0.89) | 0.73 | 0.372 (0.37-0.38) |  |  |
| LVPW | Male | 7004 | 1.12 (0.22) | 1.17 (0.19) | 0.23 | 0.146 (0.14-0.15) | <0.0001 | 0.009 |
|  | Female | 5305 | 1.05 (0.23) | 1.08 (0.19) | 0.34 | 0.137 (0.13-0.14) |  |  |
| Left Atrium | Male | 4912 | 4.02 (0.77) | 3.99 (0.7) | 0.64 | 0.332 (0.32-0.34) | <0.0001 | 0.039 |
|  | Female | 4104 | 3.63 (0.74) | 3.59 (0.66) | 0.70 | 0.293 (0.28-0.30) |  |  |
| Ascending Aorta | Male | 3904 | 3.37 (0.52) | 3.33 (0.49) | 0.19 | 0.321 (0.31-0.33) | <0.0001 | 0.032 |
|  | Female | 2809 | 3.07 (0.49) | 3.06 (0.48) | 0.25 | 0.289 (0.28-0.30) |  |  |
| Aortic root | Male | 5718 | 3.36 (0.44) | 3.4 (0.45) | 0.23 | 0.250 (0.24-0.26) | 0.04 | 0.012 |
|  | Female | 4368 | 2.96 (0.39) | 3.01 (0.4) | 0.10 | 0.238 (0.23-0.25) |  |  |
| RV Base | Male | 5289 | 3.83 (0.7) | 3.81 (0.6) | 0.55 | 0.349 (0.34-0.36) | <0.0001 | 0.02 |
|  | Female | 4143 | 3.44 (0.68) | 3.42 (0.58) | 0.56 | 0.329 (0.32-0.34) |  |  |
| Pulmonary Artery | Male | 485 | 2.1 (0.65) | 2.14 (0.46) | 0.34 | 0.401 (0.37-0.43) | 0.04 | 0.045 |
|  | Female | 550 | 2.01 (0.7) | 2.02 (0.47) | 0.37 | 0.446 (0.42-0.47) |  |  |
| IVC | Male | 1944 | 1.75 (0.49) | 1.72 (0.41) | 0.51 | 0.252 (0.24-0.26) | 0.02 | 0.019 |
|  | Female | 1363 | 1.59 (0.47) | 1.58 (0.39) | 0.52 | 0.233 (0.22-0.25) |  |  |
| **Doppler Measurements** |  |  |  |  |  |  |  |  |
| AV Vmax | Male | 13345 | 127.1 (37.36) | 136.26 (40.13) | 0.67 | 12.938 (12.65-13.23) | 0.36 | 0.19 |
|  | Female | 10686 | 134.96 (38.2) | 144.32 (40.15) | 0.74 | 12.748 (12.47-13.02) |  |  |
| TR Vmax | Male | 60209 | 246.7 (56.13) | 242.47 (65.46) | 0.6 | 26.753 (26.57-26.94) | <0.0001 | 1.69 |
|  | Female | 53545 | 253.14 (59.88) | 254.07 (69.3) | 0.7 | 25.064 (24.89-25.25) |  |  |
| MR Vmax | Male | 4586 | 425.57 (105.89) | 381.33 (127.24) | 0.45 | 55.655 (54.07-57.25) | <0.0001 | 10.44 |
|  | Female | 3739 | 443.57 (112.04) | 415.43 (134.34) | 0.65 | 45.220 (43.65-46.80) |  |  |
| Septal e' | Male | 10220 | 7.19 (2.51) | 7.3 (2.49) | 0.79 | 0.706 (0.69-0.72) | 0.54 | 0.71 |
|  | Female | 8159 | 7.3 (2.7) | 7.44 (2.64) | 0.82 | 0.714 (0.69-0.73) |  |  |
| Lateral e' | Male | 27053 | 9.37 (3.3) | 9.5 (3.16) | 0.83 | 0.839 (0.83-0.85) | 0.25 | 0.84 |
|  | Female | 21973 | 9.02 (3.37) | 9.16 (3.28) | 0.84 | 0.828 (0.81-0.84) |  |  |
| LVOT Vmax | Male | 1870 | 94.75 (33.93) | 105.3 (34.82) | 0.63 | 15.323 (14.71-15.95) | 0.05 | 1.28 |
|  | Female | 1269 | 106.12 (48.36) | 116.29 (54.37) | 0.65 | 16.599 (15.43-17.97) |  |  |
| E velocity | Male | 15517 | 83.66 (28.36) | 84.91 (28.61) | 0.81 | 8.051 (7.91-8.20) | 0.10 | 0.19 |
|  | Female | 11565 | 89.9 (30.66) | 91.76 (30.71) | 0.84 | 8.238 (8.07-8.41) |  |  |
| E/A | Male | 15519 | 1.49 (0.82) | 1.45 (0.77) | 0.76 | 0.226 (0.22-0.23) | <0.0001 | 0.03 |
|  | Female | 11565 | 1.37 (0.75) | 1.35 (0.71) | 0.79 | 0.192 (0.19-0.20) |  |  |
| TAPSE | Male | 14312 | 1.9 (0.58) | 1.87 (0.56) | 0.92 | 0.054 (0.05-0.06) | <0.0001 | 0.006 |
|  | Female | 11760 | 1.93 (0.51) | 1.91 (0.51) | 0.98 | 0.048 (0.05-0.05) |  |  |

MAE: mean absolute error, DL: Deep learning. For a detailed explanation of echocardiography parameter abbreviations, refer to Supplemental Table 1.

**Supplemental Table 6:**
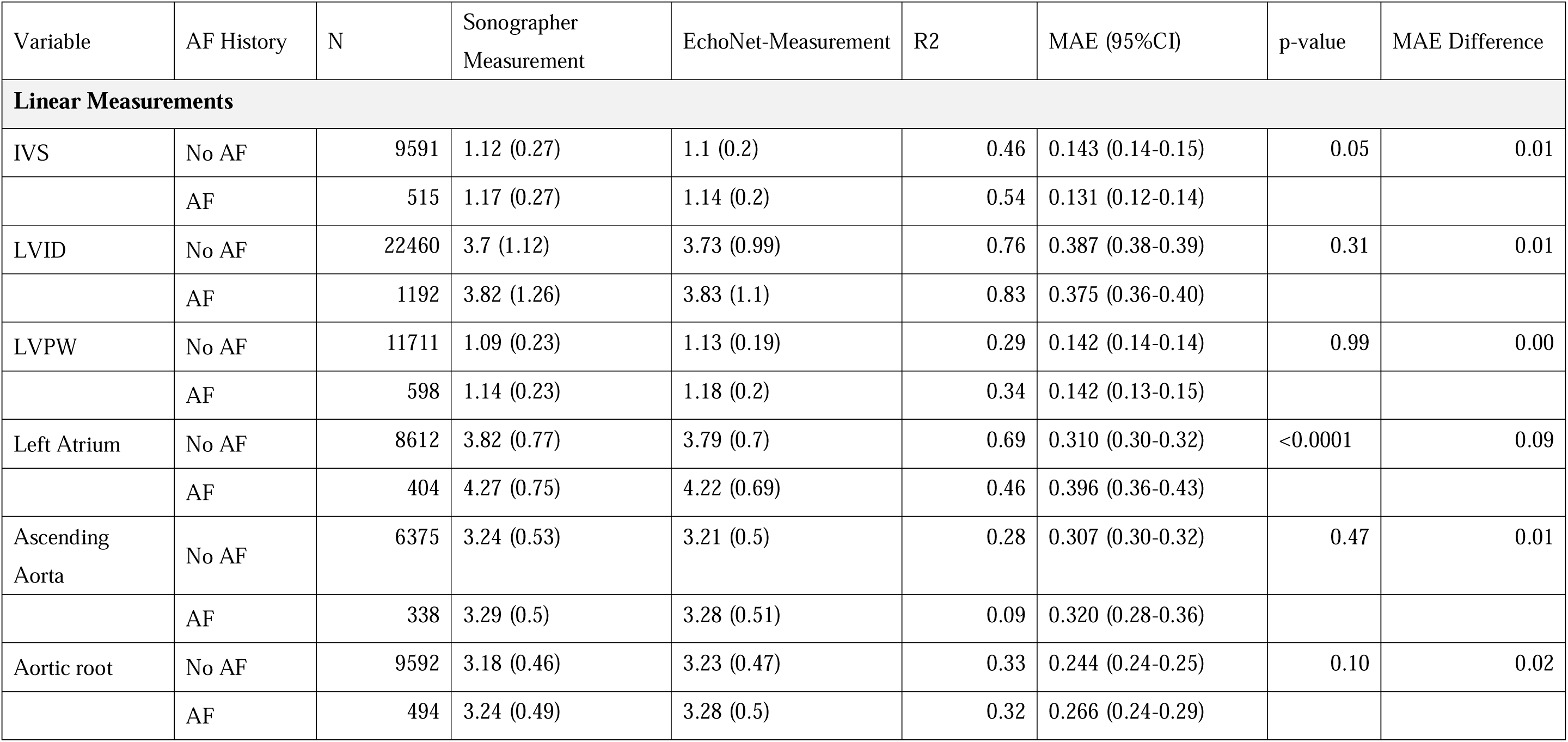
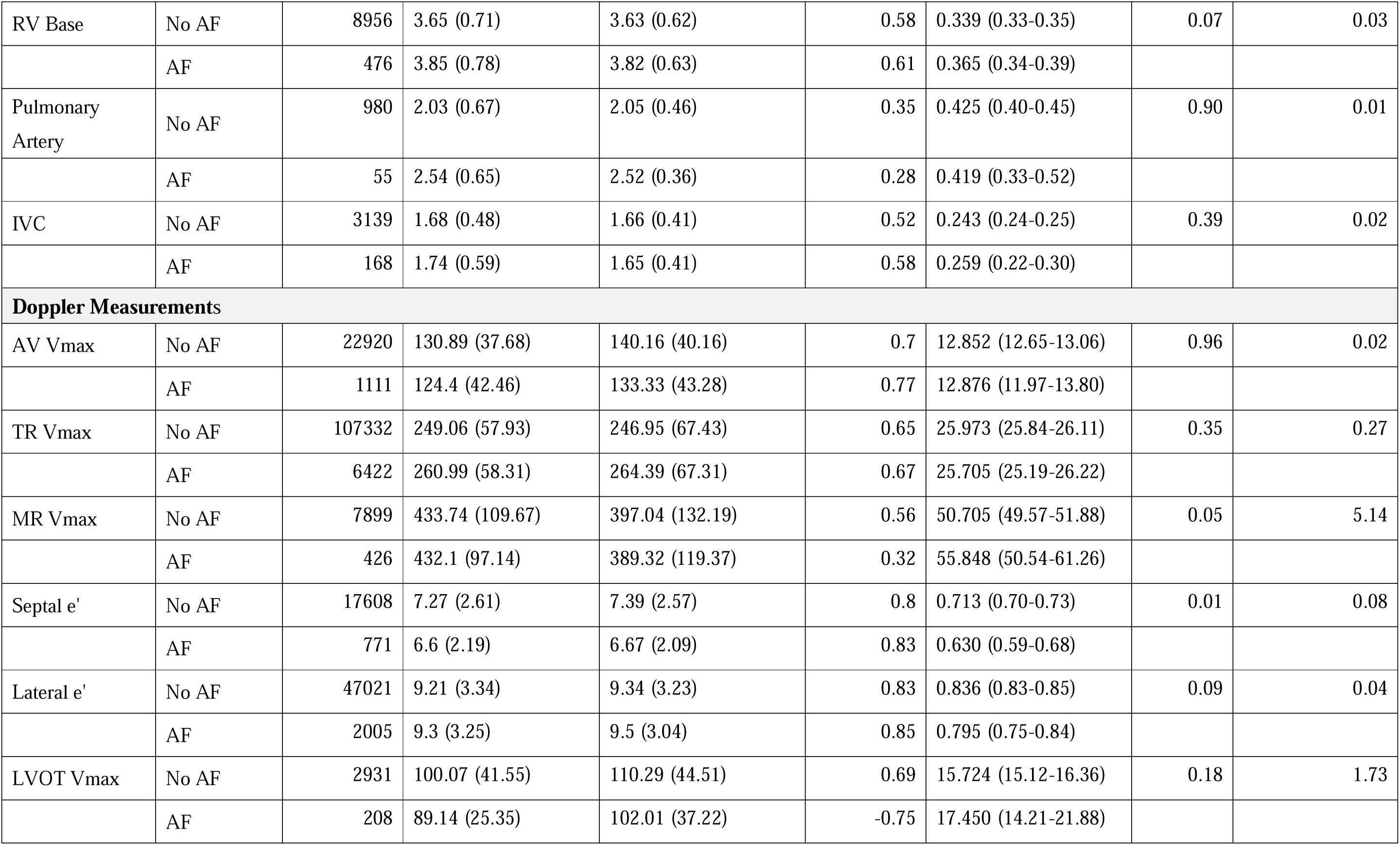
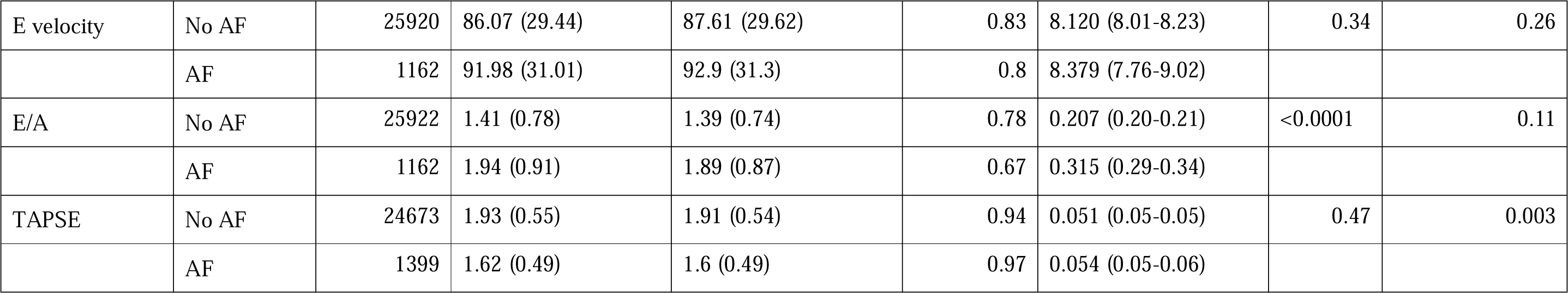
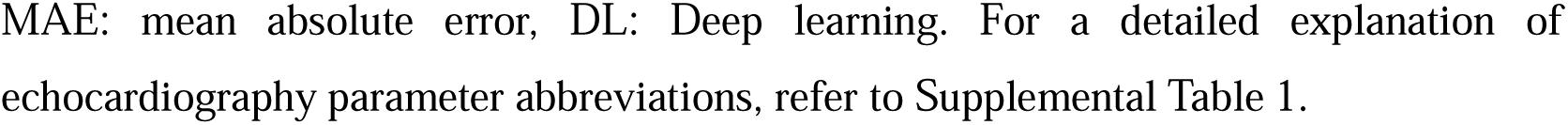
Evaluation of Deep Learning Model Performance by a History of Atrial Fibrillation (CSMC Held-Out Dataset)

| Variable | AF History | N | Sonographer Measurement | EchoNet-Measurement | R2 | MAE (95%CI) | p-value | MAE Difference |
| --- | --- | --- | --- | --- | --- | --- | --- | --- |
| **Linear Measurements** |  |  |  |  |  |  |  |  |
| IVS | No AF | 9591 | 1.12 (0.27) | 1.1 (0.2) | 0.46 | 0.143 (0.14-0.15) | 0.05 | 0.01 |
|  | AF | 515 | 1.17 (0.27) | 1.14 (0.2) | 0.54 | 0.131 (0.12-0.14) |  |  |
| LVID | No AF | 22460 | 3.7 (1.12) | 3.73 (0.99) | 0.76 | 0.387 (0.38-0.39) | 0.31 | 0.01 |
|  | AF | 1192 | 3.82 (1.26) | 3.83 (1.1) | 0.83 | 0.375 (0.36-0.40) |  |  |
| LVPW | No AF | 11711 | 1.09 (0.23) | 1.13 (0.19) | 0.29 | 0.142 (0.14-0.14) | 0.99 | 0.00 |
|  | AF | 598 | 1.14 (0.23) | 1.18 (0.2) | 0.34 | 0.142 (0.13-0.15) |  |  |
| Left Atrium | No AF | 8612 | 3.82 (0.77) | 3.79 (0.7) | 0.69 | 0.310 (0.30-0.32) | <0.0001 | 0.09 |
|  | AF | 404 | 4.27 (0.75) | 4.22 (0.69) | 0.46 | 0.396 (0.36-0.43) |  |  |
| Ascending Aorta | No AF | 6375 | 3.24 (0.53) | 3.21 (0.5) | 0.28 | 0.307 (0.30-0.32) | 0.47 | 0.01 |
|  | AF | 338 | 3.29 (0.5) | 3.28 (0.51) | 0.09 | 0.320 (0.28-0.36) |  |  |
| Aortic root | No AF | 9592 | 3.18 (0.46) | 3.23 (0.47) | 0.33 | 0.244 (0.24-0.25) | 0.10 | 0.02 |
|  | AF | 494 | 3.24 (0.49) | 3.28 (0.5) | 0.32 | 0.266 (0.24-0.29) |  |  |
| RV Base | No AF | 8956 | 3.65 (0.71) | 3.63 (0.62) | 0.58 | 0.339 (0.33-0.35) | 0.07 | 0.03 |
|  | AF | 476 | 3.85 (0.78) | 3.82 (0.63) | 0.61 | 0.365 (0.34-0.39) |  |  |
| Pulmonary Artery | No AF | 980 | 2.03 (0.67) | 2.05 (0.46) | 0.35 | 0.425 (0.40-0.45) | 0.90 | 0.01 |
|  | AF | 55 | 2.54 (0.65) | 2.52 (0.36) | 0.28 | 0.419 (0.33-0.52) |  |  |
| IVC | No AF | 3139 | 1.68 (0.48) | 1.66 (0.41) | 0.52 | 0.243 (0.24-0.25) | 0.39 | 0.02 |
|  | AF | 168 | 1.74 (0.59) | 1.65 (0.41) | 0.58 | 0.259 (0.22-0.30) |  |  |
| **Doppler Measurements** |  |  |  |  |  |  |  |  |
| AV Vmax | No AF | 22920 | 130.89 (37.68) | 140.16 (40.16) | 0.7 | 12.852 (12.65-13.06) | 0.96 | 0.02 |
|  | AF | 1111 | 124.4 (42.46) | 133.33 (43.28) | 0.77 | 12.876 (11.97-13.80) |  |  |
| TR Vmax | No AF | 107332 | 249.06 (57.93) | 246.95 (67.43) | 0.65 | 25.973 (25.84-26.11) | 0.35 | 0.27 |
|  | AF | 6422 | 260.99 (58.31) | 264.39 (67.31) | 0.67 | 25.705 (25.19-26.22) |  |  |
| MR Vmax | No AF | 7899 | 433.74 (109.67) | 397.04 (132.19) | 0.56 | 50.705 (49.57-51.88) | 0.05 | 5.14 |
|  | AF | 426 | 432.1 (97.14) | 389.32 (119.37) | 0.32 | 55.848 (50.54-61.26) |  |  |
| Septal e' | No AF | 17608 | 7.27 (2.61) | 7.39 (2.57) | 0.8 | 0.713 (0.70-0.73) | 0.01 | 0.08 |
|  | AF | 771 | 6.6 (2.19) | 6.67 (2.09) | 0.83 | 0.630 (0.59-0.68) |  |  |
| Lateral e' | No AF | 47021 | 9.21 (3.34) | 9.34 (3.23) | 0.83 | 0.836 (0.83-0.85) | 0.09 | 0.04 |
|  | AF | 2005 | 9.3 (3.25) | 9.5 (3.04) | 0.85 | 0.795 (0.75-0.84) |  |  |
| LVOT Vmax | No AF | 2931 | 100.07 (41.55) | 110.29 (44.51) | 0.69 | 15.724 (15.12-16.36) | 0.18 | 1.73 |
|  | AF | 208 | 89.14 (25.35) | 102.01 (37.22) | -0.75 | 17.450 (14.21-21.88) |  |  |
| E velocity | No AF | 25920 | 86.07 (29.44) | 87.61 (29.62) | 0.83 | 8.120 (8.01-8.23) | 0.34 | 0.26 |
|  | AF | 1162 | 91.98 (31.01) | 92.9 (31.3) | 0.8 | 8.379 (7.76-9.02) |  |  |
| E/A | No AF | 25922 | 1.41 (0.78) | 1.39 (0.74) | 0.78 | 0.207 (0.20-0.21) | <0.0001 | 0.11 |
|  | AF | 1162 | 1.94 (0.91) | 1.89 (0.87) | 0.67 | 0.315 (0.29-0.34) |  |  |
| TAPSE | No AF | 24673 | 1.93 (0.55) | 1.91 (0.54) | 0.94 | 0.051 (0.05-0.05) | 0.47 | 0.003 |
|  | AF | 1399 | 1.62 (0.49) | 1.6 (0.49) | 0.97 | 0.054 (0.05-0.06) |  |  |

MAE: mean absolute error, DL: Deep learning. For a detailed explanation of echocardiography parameter abbreviations, refer to Supplemental Table 1.

**Supplemental Table 7:**
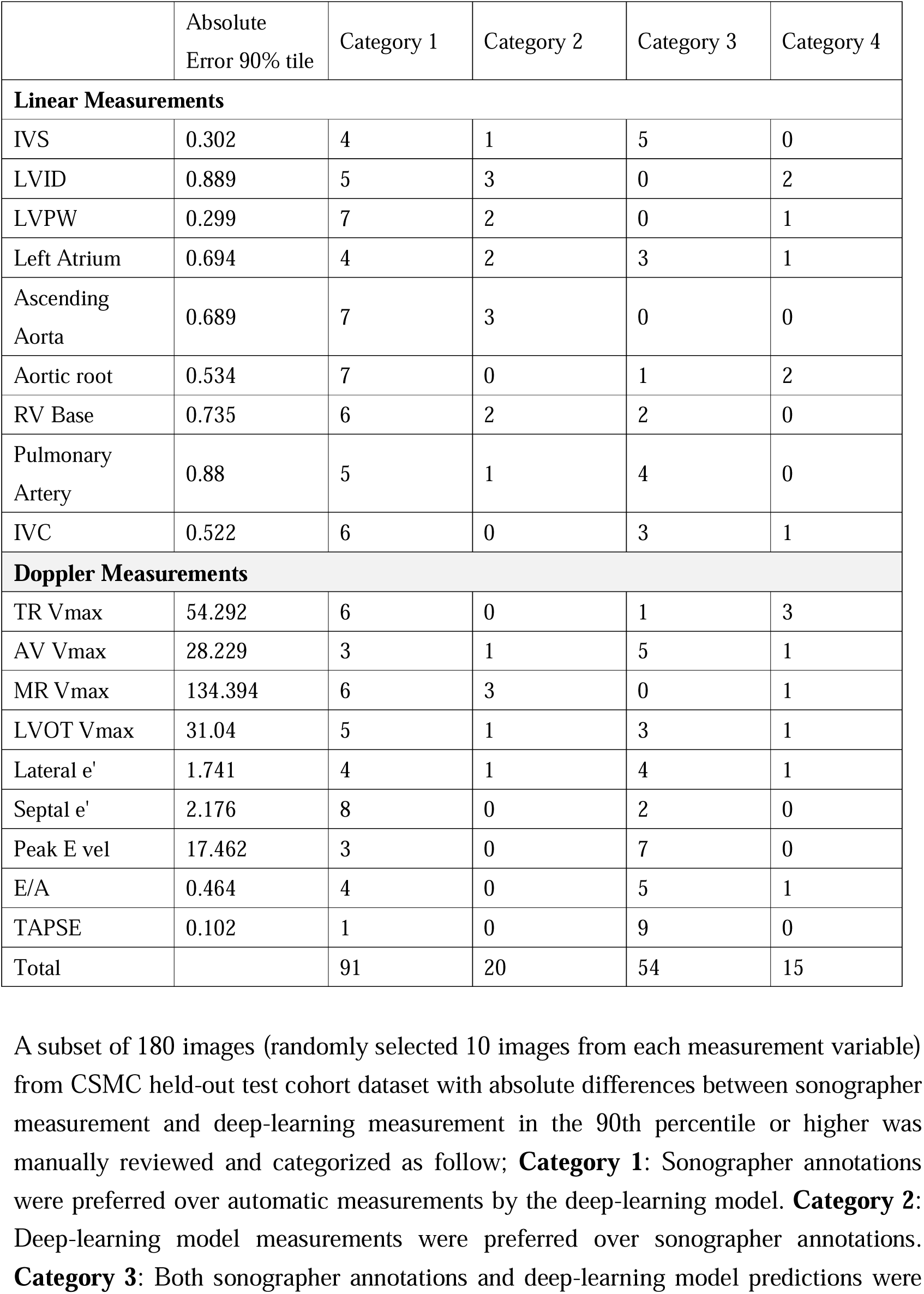
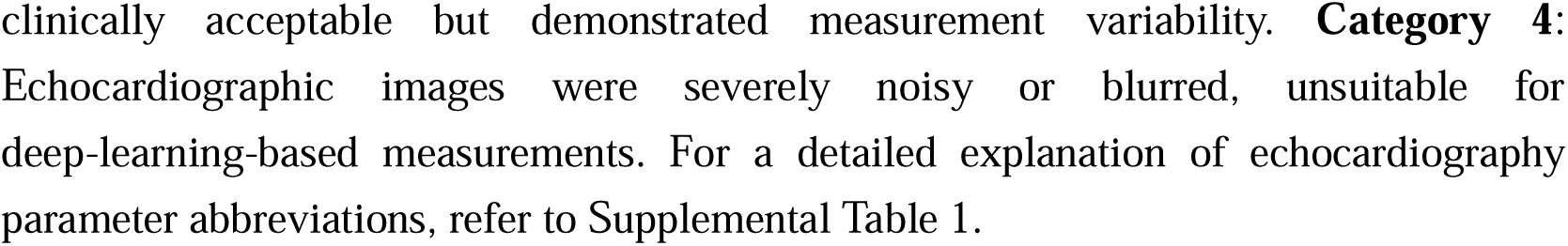
Categorization of Manually Reviewed Images with Measurement Discrepancies by Echocardiographic Parameters.

|  | Absolute<br>Error 90% tile | Category 1 | Category 2 | Category 3 | Category 4 |
| --- | --- | --- | --- | --- | --- |
| **Linear Measurements** |  |  |  |  |  |
| IVS | 0.302 | 4 | 1 | 5 | 0 |
| LVID | 0.889 | 5 | 3 | 0 | 2 |
| LVPW | 0.299 | 7 | 2 | 0 | 1 |
| Left Atrium | 0.694 | 4 | 2 | 3 | 1 |
| Ascending<br>Aorta | 0.689 | 7 | 3 | 0 | 0 |
| Aortic root | 0.534 | 7 | 0 | 1 | 2 |
| RV Base | 0.735 | 6 | 2 | 2 | 0 |
| Pulmonary<br>Artery | 0.88 | 5 | 1 | 4 | 0 |
| IVC | 0.522 | 6 | 0 | 3 | 1 |
| **Doppler Measurements** |  |  |  |  |  |
| TR Vmax | 54.292 | 6 | 0 | 1 | 3 |
| AV Vmax | 28.229 | 3 | 1 | 5 | 1 |
| MR Vmax | 134.394 | 6 | 3 | 0 | 1 |
| LVOT Vmax | 31.04 | 5 | 1 | 3 | 1 |
| Lateral e' | 1.741 | 4 | 1 | 4 | 1 |
| Septal e' | 2.176 | 8 | 0 | 2 | 0 |
| Peak E vel | 17.462 | 3 | 0 | 7 | 0 |
| E/A | 0.464 | 4 | 0 | 5 | 1 |
| TAPSE | 0.102 | 1 | 0 | 9 | 0 |
| Total |  | 91 | 20 | 54 | 15 |

A subset of 180 images (randomly selected 10 images from each measurement variable) from CSMC held-out test cohort dataset with absolute differences between sonographer measurement and deep-learning measurement in the 90th percentile or higher was manually reviewed and categorized as follow; **Category 1:** Sonographer annotations were preferred over automatic measurements by the deep-learning model. **Category 2:** Deep-learning model measurements were preferred over sonographer annotations. **Category 3:** Both sonographer annotations and deep-learning model predictions were

**Supplemental Figure 1:**
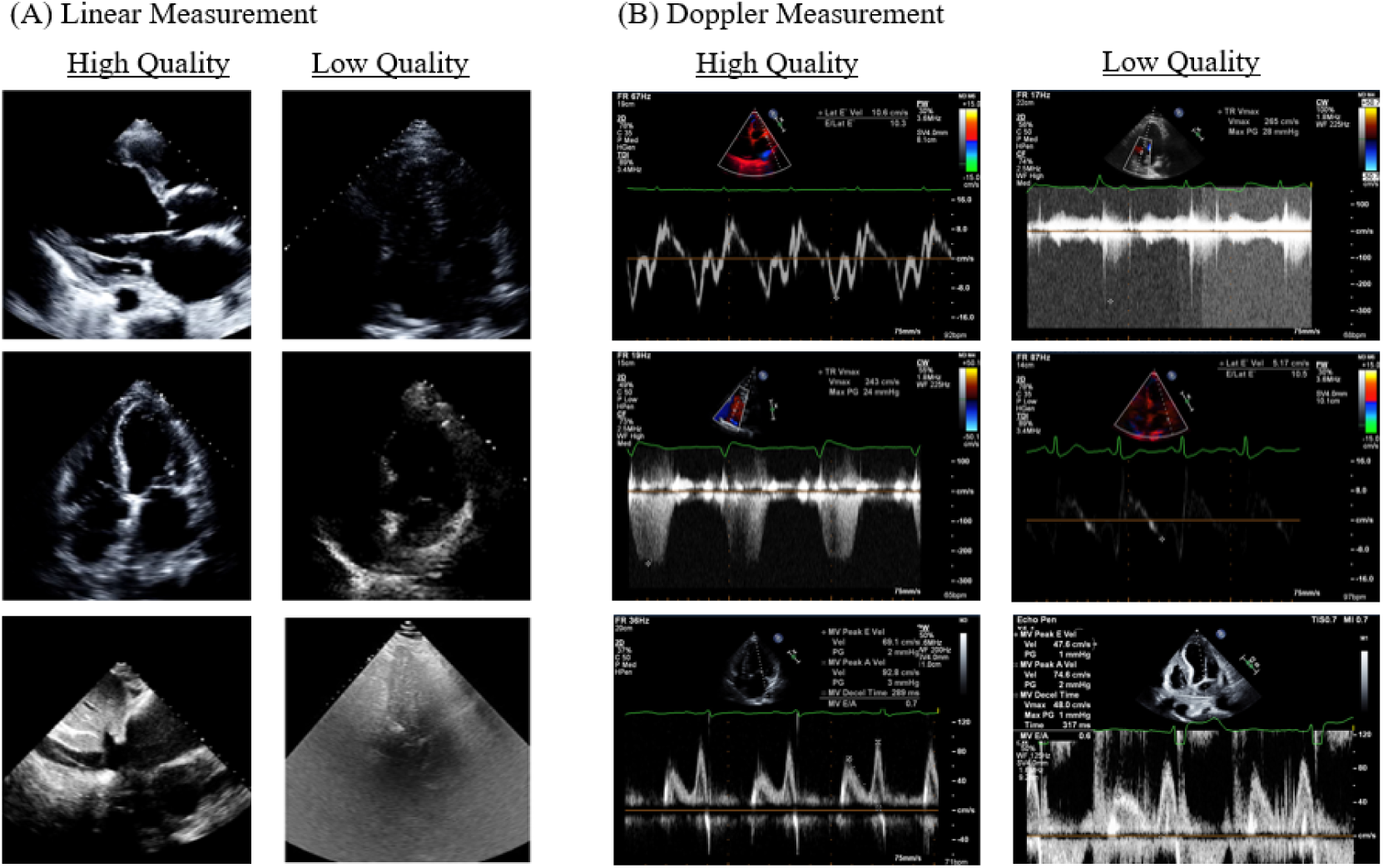
Representative images of quality control model dataset. Representative images demonstrating variations in image quality within the quality control model dataset. High-quality images (left) and low-quality images (right) with blur or noise in the linear measurement model (A) and Doppler imaging models (B).

**Supplemental Figure 2:**
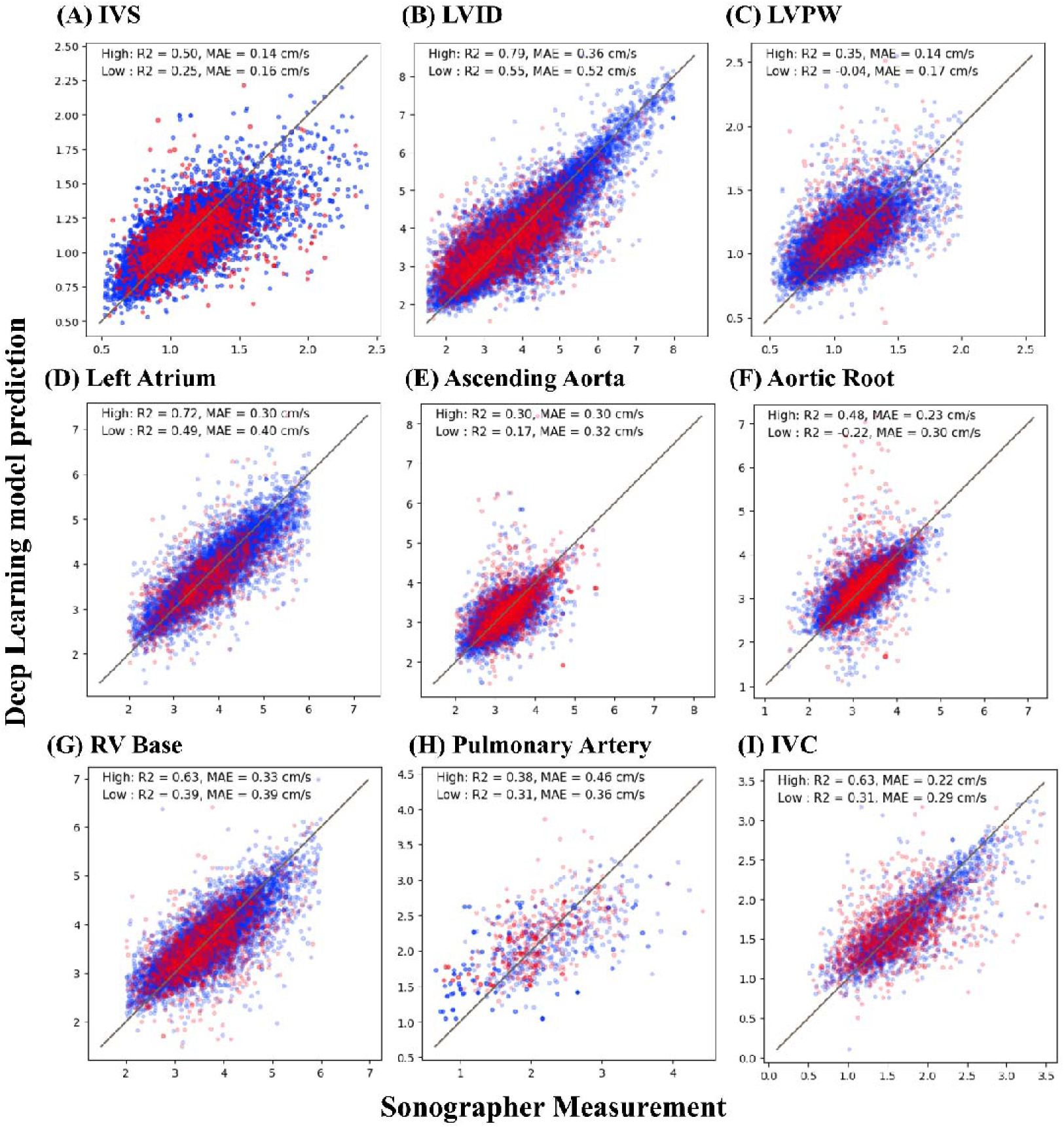
Correlation between Deep Learning Model and Sonographer Annotations for Echocardiographic Linear Measurements of B-mode Classified by Predicted Image-Quality. Scatterplot between deep learning model predictions and sonographer annotations for various echocardiographic linear measurements, classified by predicted image quality. Each plot represents a specific measurement, with the x-axis showing the values annotated by the sonographer and the y-axis showing the values predicted by the Deep Learning model. Blue dots indicate higher quality images predicted by image-quality model, and red dots indicate lower quality images predicted by the same model. The coefficient of determination (R2) and mean absolute error (MAE). For a detailed explanation of echocardiography parameter abbreviations, refer to Supplemental Table 1.

**Supplemental Figure 3:**
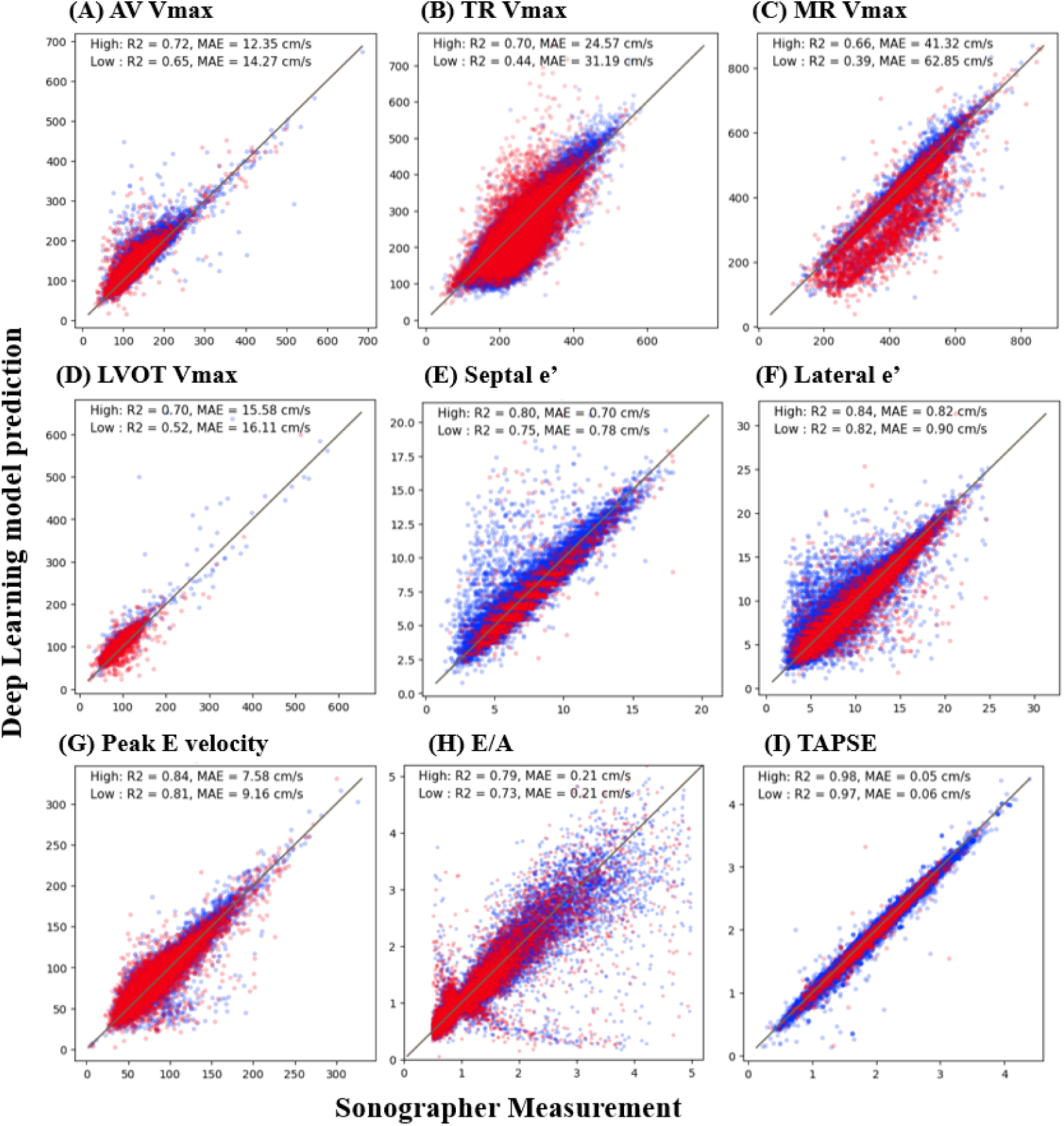
Correlation between Deep Learning Model and Sonographer Annotations for Echocardiographic Doppler Measurements Classified by Predicted Image-Quality. Scatterplot between deep learning model predictions and sonographer annotations for various echocardiographic Doppler measurements, classified by predicted image quality. Each plot represents a specific measurement, with the x-axis showing the values annotated by the sonographer and the y-axis showing the values predicted by the Deep Learning model. Blue indicates higher quality images predicted by image-quality model, and red indicates lower quality images predicted by the same model. The coefficient of determination (R2) and mean absolute error (MAE). For a detailed explanation of echocardiography parameter abbreviations, refer to Supplemental Table 1.

**Supplemental Figure 4:**
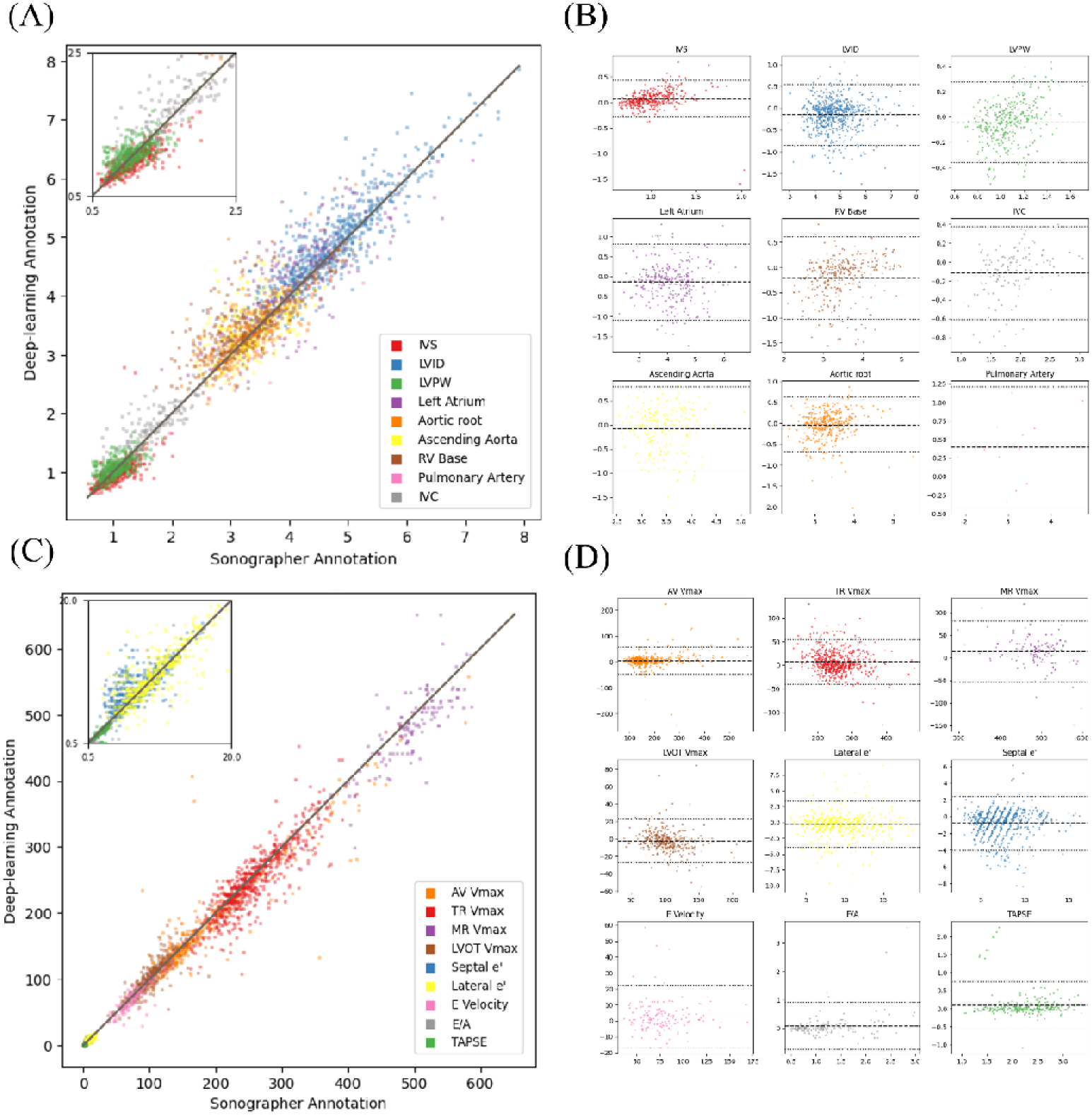
Model Performance and Agreement of EchoNet-Measurement in the external test dataset with Study-level ground truth value. (A and C) Scatter plots comparing deep learning model predictions with sonographer annotations for linear measurements (A) and Doppler echocardiography parameters (C). (B and D) Bland-Altman plots for each parameter in the linear measurement group (B) and Doppler echocardiography parameters (D). All metrics including coefficient of determination (R2), intraclass correlation coefficients (ICC), mean absolute error (MAE), bias and limits of agreement are described in **Supplemental Table 3** (Stanford Health Care dataset with study-level ground truth measurement value), respectively. Refer to Table 1 for a detailed explanation of echocardiography parameter abbreviations, Figure 2 for the explanation of figure legend and Bland-Altman plot. In Bland-Altman Plot (B and D), gray and black lines indicate bias and limits of agreement.

**Supplemental Figure 5:**
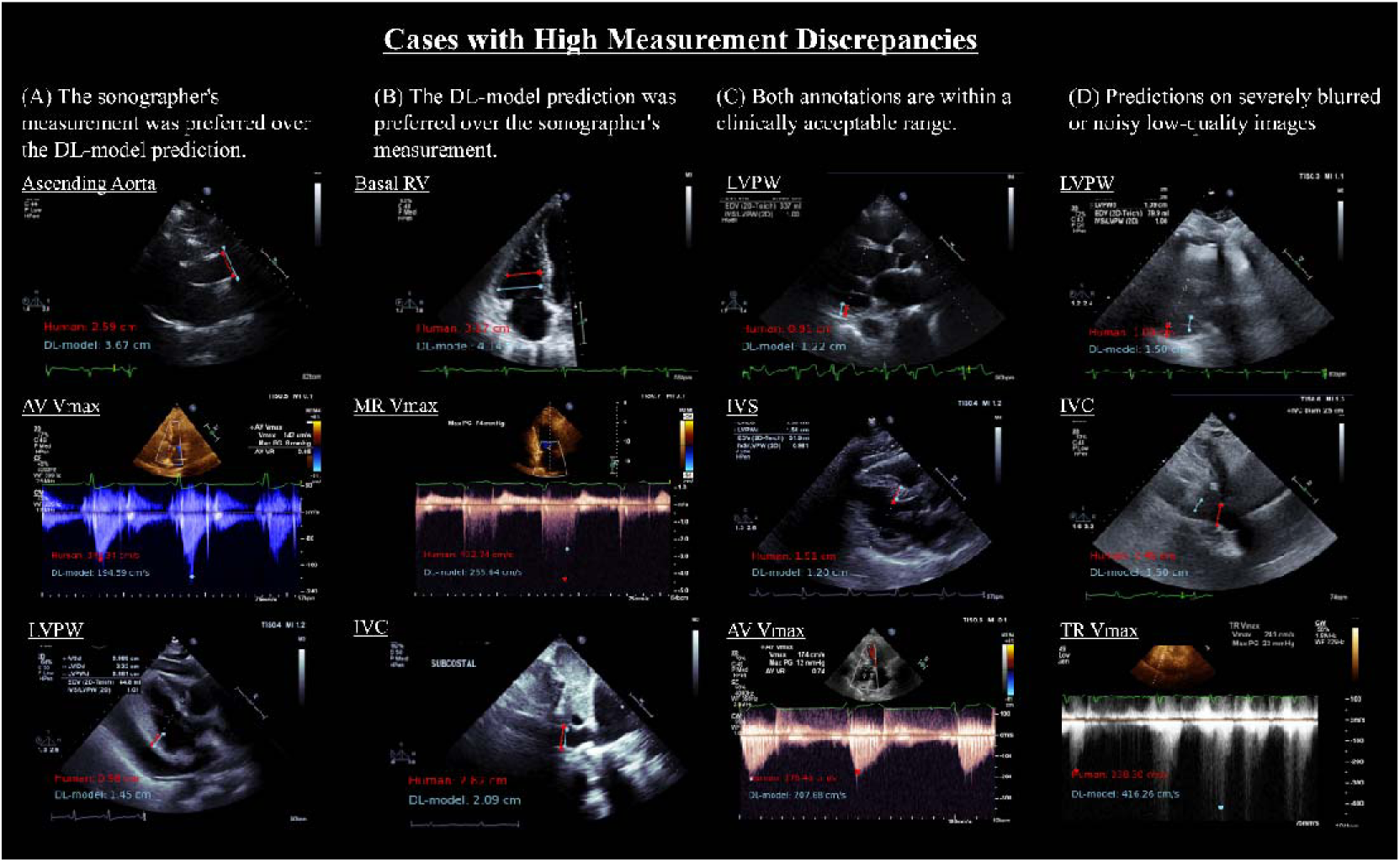
Representative Cases of High Measurement Discrepancies Analysis in Echocardiographic Measurements. Representative echocardiography images of high measurement discrepancies analysis. Reviewed images were categorized into four categories; (A) cases where the reviewer considered that the sonographer’s measurement was preferred over the deep-learning (DL) model prediction, (B) cases where the DL model’s prediction was preferred over the sonographer’s measurement, (C) cases where both annotations (sonographer and DL model) were within a clinically acceptable range despite measurement variability, (D) cases where the echocardiographic images were severely noisy or blurred, considered inappropriate images for the DL-model input. Red texts and dots indicate sonographer measurements, and blue texts and dot show the DL model’s measurements. For a detailed explanation of echocardiography parameter abbreviations, refer to Supplemental Table 1.

**Supplemental Video 1**: Beat-to-beat Linear Measurement Parameters Prediction Legend: This video demonstrates (A through I) the deep learning model’s beat-to-beat predictions for multiple linear measurement parameters across echocardiographic images. For a detailed explanation of echocardiography parameter abbreviations, refer to Supplemental Table 1.

**Supplemental Video 2:** Demo for High-Throughput TR Vmax Measurement and Annotation Pipeline for CW Doppler Waveform Prediction

Legend: This interface developed using Gradio demonstrates a process for uploading multiple DICOM images containing TR Vmax measurements and performing high-throughput annotations using an automated pipeline. Users can upload multiple DICOM files and these are then processed to annotate the predicted points and calculate the TR Vmax values. The outputs including annotated Doppler images and extracted TR Vmax values are displayed in a results table and can be exported as a CSV file.

## Supplemental Methods

For the development of the quality-control model for the linear measurement group, a video-based convolutional neural network (R2+1D) was used with standardized input videos of 112 × 112 pixel for linear measurement group. For the Doppler image quality-control model, an image-based model (DenseNet) was employed with 426 × 1024 still image inputs. A dataset of 31,506 manually curated videos (9.8% were classified as poor quality videos) from 21,035 patients and 29,232 still images (21.5% were classified as poor images or noisy images) from 20,951 patients in CSMC was split 8:1:1 ratio by patient medical record number and all videos and images were classified as low-quality or high-quality images (representative ground truth images in **Supplemental Figure 2**). Image quality was evaluated by two cardiologists (Y.S and C.B.R). The model was trained to minimize binary cross-entropy loss using the AdamW optimizer. The initial learning rate was set to 1×10-5, with a batch size of 10 for the video model and 24 for the image model, over the course of 50 epochs with an early stopping setting. Model performance was evaluated in the held-out dataset, achieving an AUROC of 0.984 in the linear measurement group and an AUROC of 0.912 in the Doppler measurement group.

Further, a classification model was developed using a similar procedure to exclude cases where annotated point was plotted on a’ during septal or lateral e’ prediction. Datasets were manually reviewed and finally datasets annotated with only e’ (n=7962) or only a’ (n=2128) were prepared. A classification model was then built using DenseNet and abovementioned hyperparameters and image sizes. This model achieved an AUROC of 0.965 on a test dataset split by patient in an 8:1:1 ratio, and the optimal cutoff value was determined using the Youden Index. Following the inference of septal e’ and lateral e’ prediction, data plotted as a’ were removed by passing them through this model.

## Author contributions

Concept and design: YS, DO,

Code and programming: YS, HI, MV, MC, JR, BH, DO,

Data acquisition, analysis, or interpretation of data: YS, VY, CBR, DO, PC

Drafting and Critical revision of the manuscript: YS, DO

Statistical analysis: YS, DO, HI

Obtained funding: DO

Supervision: DO

